# Lower levels of household transmission of SARS-CoV-2 VOC Omicron compared to Wild-type: an interplay between transmissibility and immune status

**DOI:** 10.1101/2024.07.16.24310515

**Authors:** AMAM Winkel, E Kozanli, ME Haverkort, SM Euser, JGC Sluiter-Post, R Mariman, A. Vogelzang, J. de Bakker, C.R. Lap, MA van Houten, D Eggink, SFL van Lelyveld

**Author notes:** Corresponding authors and shared last authors: D Eggink,. SFL van Lelyveld. shared first authors (AMAM Winkel, E Kozanli).

## Abstract

**Background:** Knowledge of SARS-CoV-2 household transmission dynamics guides infection control and vaccination measures. This household cohort study prospectively assessed the impact of both the Omicron BA.2 variant and immunity on household transmission using dense saliva sampling and sequence analysis.

**Methods:** Households consisting of a PCR-confirmed index and at least two household members were enrolled in March and April 2022 during the Omicron BA.2 wave in the Netherlands. SARS-CoV-2 PCR was performed on ten consecutive saliva samples. Serum-antibodies were measured at baseline and day 42. Whole genome sequencing was performed for phylogenetic analysis, followed by sensitivity analysis, to correct for multiple household introductions and index definition. Results were compared with the identical, early-pandemic and pre- immunisation predecessor study.

**Results:** Sixty-seven households were included, consisting of 241 individuals (median age 33.0 years). Maximum household Secondary Attack Rate (SAR) was 59.7%, per-person SAR 41.5%. Underage indexes were more likely to transmit. Transmission was negatively affected by household members’ immunity. Phylogenetic analysis showed multiple introductions in four households. Sensitivity analysis resulted in a minimal household SAR of 51.0% and per-person SAR of 28.5%.

**Conclusions:** The Omicron BA.2 variant is highly transmissible within households. Nevertheless, transmission rates are lower than our pre-immunisation reported ancestral rates: household SAR decreased from 88.2% to 59.7%, per-person SAR from 64.3% to 41.5%, implying immunity reduces transmission, even for a more transmittable variant. Regardless of immune status, children have a crucial role in Omicron household transmission. Intensive sampling and phylogenetic analysis are essential to correctly calculate transmission rate, especially in times of minimal behavioural restrictions.

## 1. Introduction

Since the first identification in December 2019, severe acute respiratory syndrome coronavirus (SARS-CoV-2) has quickly spread worldwide, causing a global pandemic characterised by the continuous emergence of new variants.^1^ The SARS-CoV-2 variant B.1.1.529, referred to as the Omicron variant of concern (VOC), was first reported in November 2021 and has become the most prevalent variant globally.^1^ This variant has diversified into various Omicron sublineages. Initially, five sublineages were identified, called BA.1-BA.5.^2^ BA.1 was introduced in the Netherlands in November 2021 and became the dominant variant within several weeks. This sublineage was rapidly replaced by BA.2, which was dominant in the Netherlands during this study in March and April 2022, followed by the dominance of BA.5 in June 2022.^3^ The Omicron VOC sublineages BA.1-BA.5 have been reported to be more infectious than previous variants.^4^ Moreover, Omicron is less sensitive to neutralising antibodies induced by vaccine or natural infection with a previous variant.^5^ These characteristics of Omicron VOCs facilitate infections in immunized individuals – even in those who received a booster vaccination, keeping SARS-CoV-2 highly transmissible.^6^

Households are of significant importance for SARS-CoV-2 transmission due to proximity and duration of exposure.^7^ Estimated SARS-CoV-2 household transmission rates vary between 9.0% and 88.2% for different variants.^8,9^ The highest per-person Secondary Attack Rates (SARs) have been reported by prospective household studies using frequent sampling.^10,11^ Likewise, our initial early-pandemic prospective cohort study using frequent saliva sampling found very high household transmission rates (88.2%).^9^ However, this study was performed between October and December 2020, when the ancestral variant was still dominating (Nextclade 20A, 20B, 20E (EU1)), and the national COVID-19 vaccination program had not yet been implemented. By March 2022, 84.9% of people in the Netherlands aged 12 and over had received the primary vaccination series, and 58.1% had received a booster vaccination.^12^ Furthermore, 60% of the Dutch population was estimated to have suffered a previous infection, resulting in 95% of the population having SARS-CoV-2 serum-antibodies.^13^ It is therefore highly likely that SARS-CoV-2 Omicron household transmission dynamics within an immunized population differ from the results described in the initial study and other pre-Omicron performed household studies.

Prospective data regarding household transmission in the Omicron era are scarce. Most published studies extracted transmission data from register data,^14–17^ or contact tracing programs,^18^ This strategy misses most asymptomatic and mild cases since SARS-CoV-2 testing is mostly performed in case of respiratory symptoms. Since infection with Omicron often leads to milder symptoms than previous variants, reported transmission rates are possibly underestimated.^19^ In addition, phylogenetic analysis, essential for the exclusion of multiple introductions from outside the household, is seldom performed. The net impact of the Omicron BA.2 VOC on household transmission among vaccinated and/or previous infected household members is of interest, since it could have implications for infection control and vaccination strategies.

The present study aimed to prospectively assess household transmission dynamics of Omicron BA.2 VOC and factors associated with transmission on household, index case, and household member levels using dense saliva sampling and phylogenetic analysis. The identical study design of both this and the initial study offered a unique opportunity to compare household transmission data between different pandemic phases.

## 2. Methods

### 2.1. Study design and participants

In this prospective cohort study, largely identical to the initial early-pandemic household transmission study,^9^ households were recruited in March and April 2022, during the SARS-CoV-2 Omicron (B.1.1.529) wave in the Netherlands, when Omicron sublineage BA.2 was dominant.

Index cases were recruited at the Public Health Services Kennemerland, the Netherlands, when testing for SARS-CoV-2 through reversed transcription polymerase chain reaction (RT-PCR) on a combined nasopharyngeal swab (NPS) and oropharyngeal swab (OPS). The indexes and their households were eligible if (1) the index was 65 years or younger and had a SARS-CoV-2 positive RT-PCR maximal 72 hours before study inclusion, and (2) at least two additional household members not diagnosed with SARS-CoV-2 in the previous eight weeks were willing to participate in the study. Indexes could be either symptomatic or asymptomatic. A household was considered to have two indexes if both participants tested positive for SARS-CoV-2 at the same moment and symptoms started simultaneously. For detailed case definitions see Supplementary Methods.

All participants provided written informed consent. The Medical Ethical Committee of the Amsterdam University Medical Centre, The Netherlands, reviewed and approved this study (reference number 2022.0073).

### 2.2. Measurements

#### Sample collection

Saliva samples were obtained by self-sampling at home on days 1, 3, 5, 7, 10, 14, 21, 28, 35, and 42, with the day of inclusion as day 1. On days 1 and 42, capillary blood samples were collected by the research team during a home visit (Figure I). The sampling scheme was identical for index cases and household members. See Supplementary Materials for details.

**Figure I.**
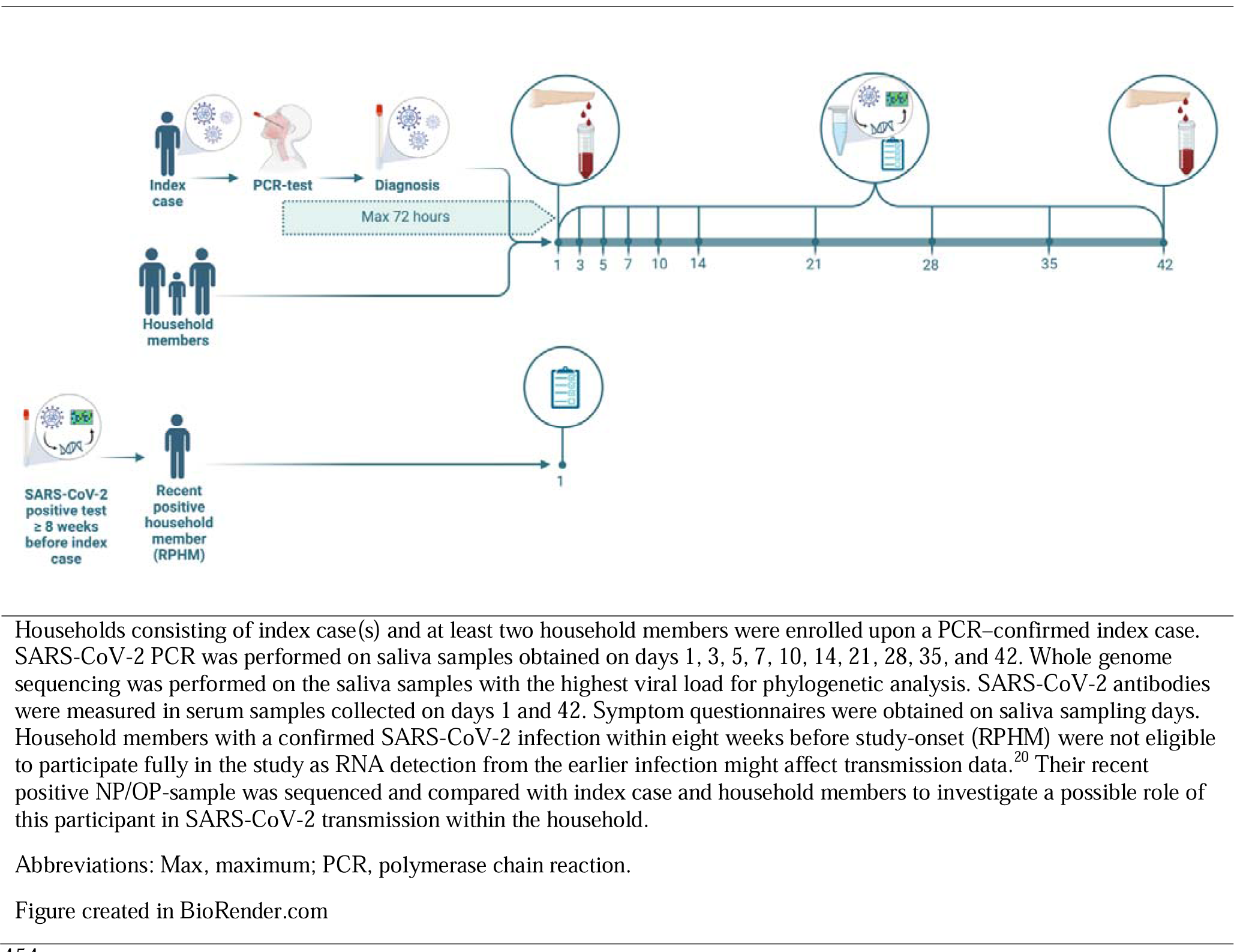
Study design.

#### Questionnaires

An online baseline questionnaire obtained on day 1 contained questions on household composition, household characteristics, medical history, previous SARS-CoV-2 infection(s), and SARS-CoV-2 vaccination status. Online symptomatology questionnaires were sent to the participants each night before the pre-defined time points for saliva collection.

#### Recently SARS-CoV-2 positive household members (RPHM)

Household members with a confirmed SARS-CoV-2 infection (by either RT-PCR or rapid antigen self-test) up to eight weeks before study onset were defined as recently SARS-CoV-2 positive household members (RPHMs).

RPHMs were not eligible to fully participate in the study as RNA detection from the earlier infection might affect transmission data.^20^ However, these participants could conceivably be the actual index of the household. To investigate a possible role of RPHMs in SARS-CoV-2 transmission within the household, their NP/OP- samples were requested from the Public Health Services and sequenced. Additionally, these household members completed a questionnaire regarding their recent SARS-CoV-2 infection, including test date, symptom start date, duration and severity of symptoms.

### 2.3. Definitions

Infection of household members during study follow-up (secondary cases) was defined as SARS-CoV-2 positive RT-PCR in the saliva of at least one of the ten time points and/or a >5-fold rise of SARS-CoV-2 specific antibodies in serum between day 1 and day 42. Household transmission was expressed in the household secondary attack rate (SAR) and per-person SAR. The household SAR, defined as SARS-CoV-2 infection of at least one household member during study follow-up, was calculated by dividing households with transmission by the total number of households. The per-person SAR, defined as the risk of household members to get infected, was calculated by dividing the number of secondary cases by the number of participating household members. Participants were divided based on self-reported immunity status into naive, previous infection only, previous vaccination only and hybrid immunity (both SARS-CoV-2 vaccination and previous infection).

### 2.4. Molecular diagnostics and serology

The methodology for molecular diagnostics and serology analysis is similar to our initial early-pandemic study and is described in detail in the Supplementary Materials.^9^ Sera of all individuals were tested for IgG-type antibodies against SARS-CoV-2 antigens. Protein microarray was used to quantify fluorescent signal of a duplicate 3-fold serial dilutions starting at 1:20.

In saliva, cycle threshold (Ct)-values were determined using RT-PCR for the presence of SARS-CoV-2. Subsequently, RT-PCR-positive samples of each participant with the lowest Ct-value were sequenced in amplicon-based SARS-CoV-2 sequencing for phylogenetic analysis (see phylogenetic analysis).^21^ The SARS- CoV-2 sequences from the participants are compared through a phylogenetic analysis. A maximum likelihood phylogenetic tree is built to display clustering of sequences (Figure II). The proximity of sequences of household members distinguishes actual transmission within a household from an external introduction. Based on this analysis, we could correct for external introductions in our sensitivity analysis to calculate a minimal SAR.

**Figure II.**
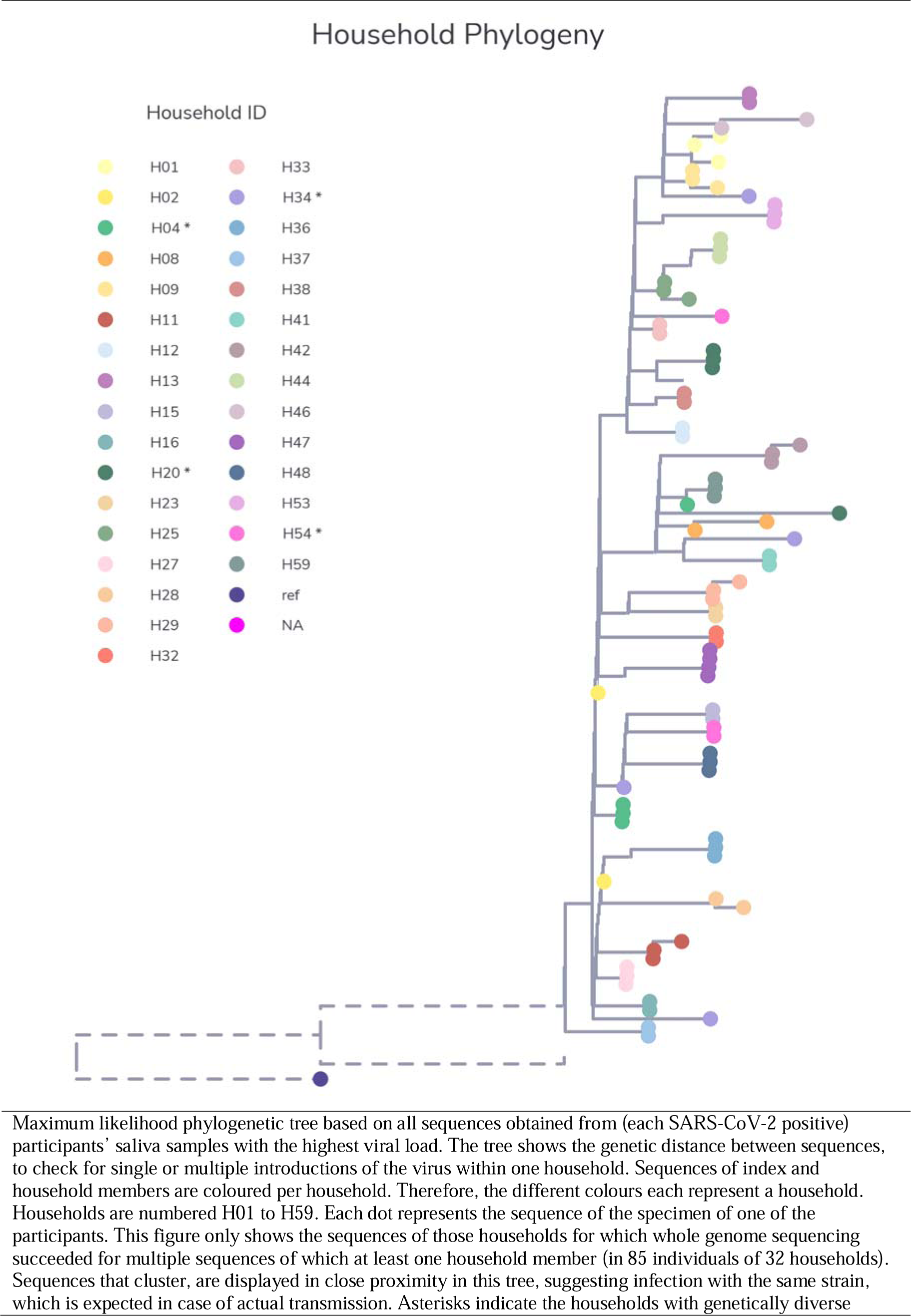
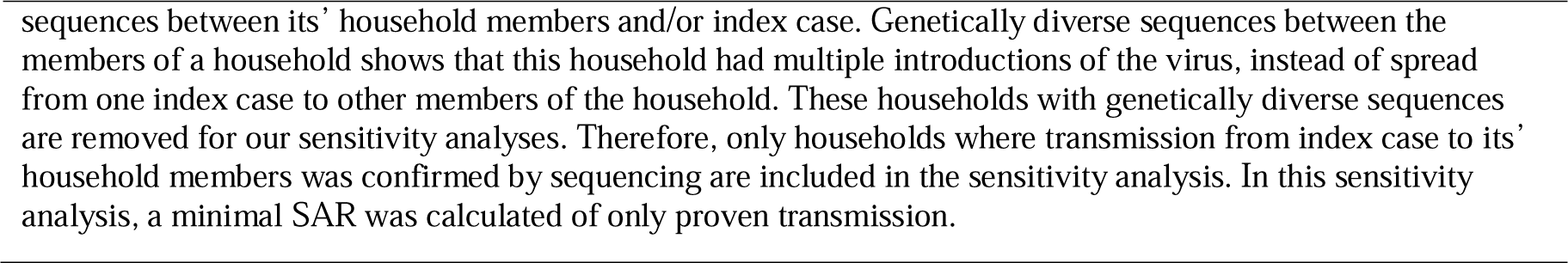
Phylogenetic analysis of SARS-CoV-2 sequences within households.

### 2.5. Statistical methods

Logistic regression was used to a) compare characteristics of households and to assess the association between household characteristics and household transmission (table II), b) compare characteristics of index cases and to assess the association between index case characteristics and transmission to its’ household members (transmission probability) (table III), c) compare characteristics of household members and to assess the association between household member characteristics and secondary infection of household members (Table IV).

**Table I.**
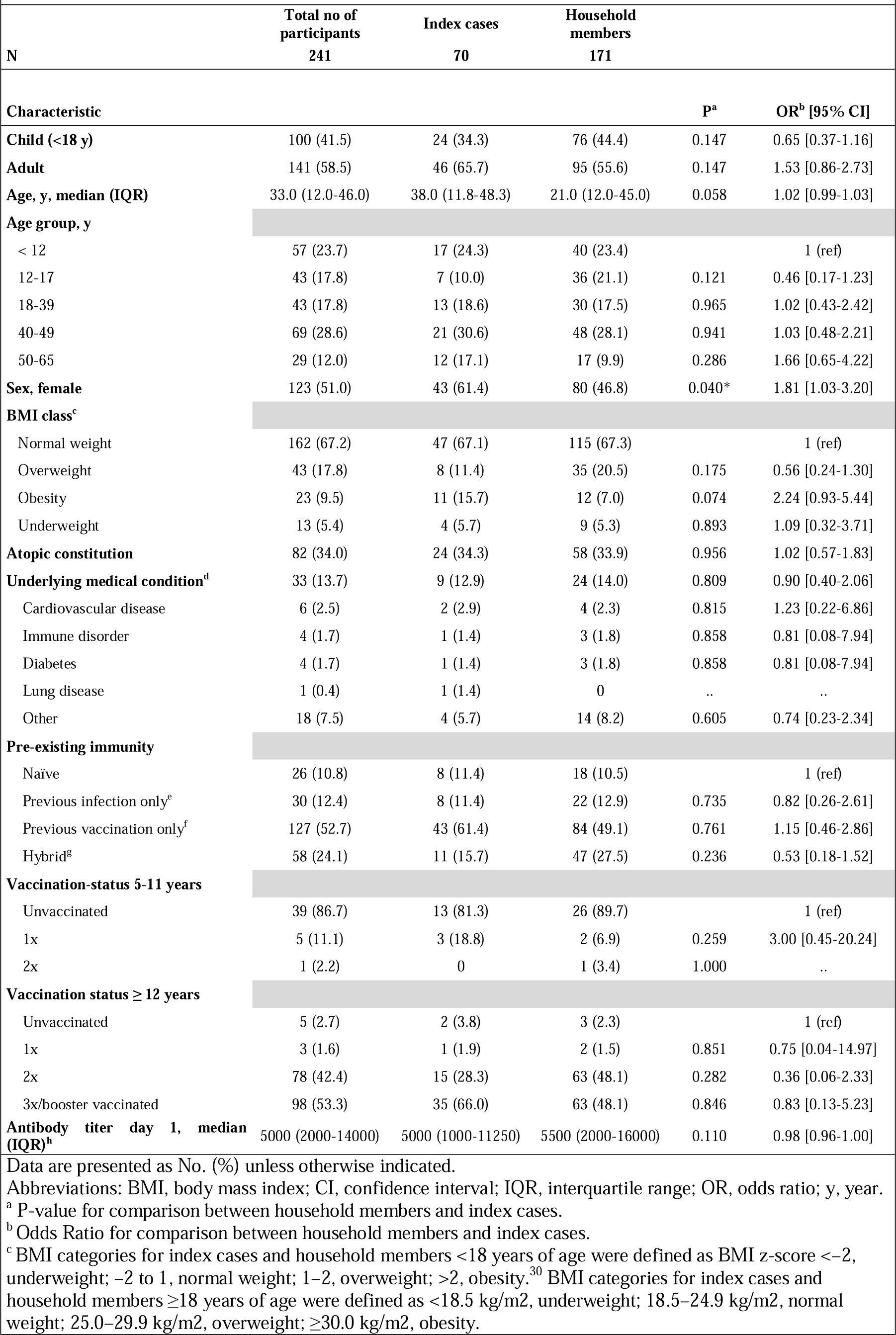
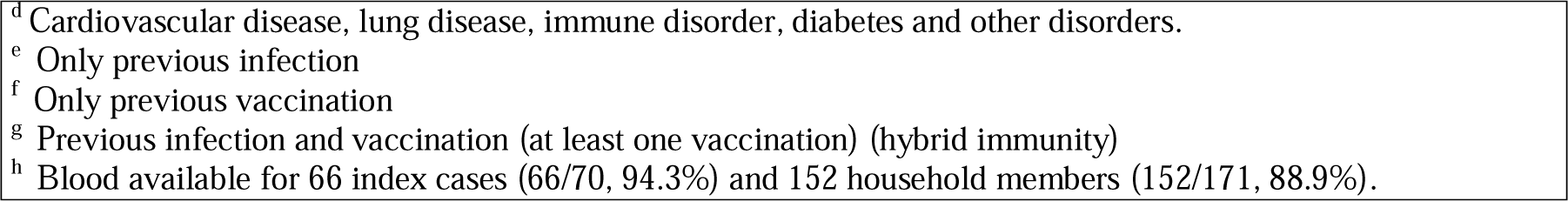
Baseline characteristics of study participants (N=241)

| Table I – Baseline characteristics of study participants (N=241) |  |  |  |  |  |
| --- | --- | --- | --- | --- | --- |
| N | Total no of participants<br>241 | Index cases<br>70 | Household members<br>171 |  |  |
| Characteristic |  |  |  | P<sup>a</sup> | OR<sup>b</sup> [95% CI] |
| **Child (<18 y)** | 100 (41.5) | 24 (34.3) | 76 (44.4) | 0.147 | 0.65 [0.37-1.16] |
| **Adult** | 141 (58.5) | 46 (65.7) | 95 (55.6) | 0.147 | 1.53 [0.86-2.73] |
| **Age, y, median (IQR)** | 33.0 (12.0-46.0) | 38.0 (11.8-48.3) | 21.0 (12.0-45.0) | 0.058 | 1.02 [0.99-1.03] |
| **Age group, y** |  |  |  |  |  |
| < 12 | 57 (23.7) | 17 (24.3) | 40 (23.4) |  | 1 (ref) |
| 12-17 | 43 (17.8) | 7 (10.0) | 36 (21.1) | 0.121 | 0.46 [0.17-1.23] |
| 18-39 | 43 (17.8) | 13 (18.6) | 30 (17.5) | 0.965 | 1.02 [0.43-2.42] |
| 40-49 | 69 (28.6) | 21 (30.6) | 48 (28.1) | 0.941 | 1.03 [0.48-2.21] |
| 50-65 | 29 (12.0) | 12 (17.1) | 17 (9.9) | 0.286 | 1.66 [0.65-4.22] |
| **Sex, female** | 123 (51.0) | 43 (61.4) | 80 (46.8) | 0.040* | 1.81 [1.03-3.20] |
| **BMI class<sup>c</sup>** |  |  |  |  |  |
| Normal weight | 162 (67.2) | 47 (67.1) | 115 (67.3) |  | 1 (ref) |
| Overweight | 43 (17.8) | 8 (11.4) | 35 (20.5) | 0.175 | 0.56 [0.24-1.30] |
| Obesity | 23 (9.5) | 11 (15.7) | 12 (7.0) | 0.074 | 2.24 [0.93-5.44] |
| Underweight | 13 (5.4) | 4 (5.7) | 9 (5.3) | 0.893 | 1.09 [0.32-3.71] |
| **Atopic constitution** | 82 (34.0) | 24 (34.3) | 58 (33.9) | 0.956 | 1.02 [0.57-1.83] |
| **Underlying medical condition<sup>d</sup>** | 33 (13.7) | 9 (12.9) | 24 (14.0) | 0.809 | 0.90 [0.40-2.06] |
| Cardiovascular disease | 6 (2.5) | 2 (2.9) | 4 (2.3) | 0.815 | 1.23 [0.22-6.86] |
| Immune disorder | 4 (1.7) | 1 (1.4) | 3 (1.8) | 0.858 | 0.81 [0.08-7.94] |
| Diabetes | 4 (1.7) | 1 (1.4) | 3 (1.8) | 0.858 | 0.81 [0.08-7.94] |
| Lung disease | 1 (0.4) | 1 (1.4) | 0 | .. | .. |
| Other | 18 (7.5) | 4 (5.7) | 14 (8.2) | 0.605 | 0.74 [0.23-2.34] |
| **Pre-existing immunity** |  |  |  |  |  |
| Naïve | 26 (10.8) | 8 (11.4) | 18 (10.5) |  | 1 (ref) |
| Previous infection only<sup>e</sup> | 30 (12.4) | 8 (11.4) | 22 (12.9) | 0.735 | 0.82 [0.26-2.61] |
| Previous vaccination only<sup>f</sup> | 127 (52.7) | 43 (61.4) | 84 (49.1) | 0.761 | 1.15 [0.46-2.86] |
| Hybrid<sup>g</sup> | 58 (24.1) | 11 (15.7) | 47 (27.5) | 0.236 | 0.53 [0.18-1.52] |
| **Vaccination-status 5-11 years** |  |  |  |  |  |
| Unvaccinated | 39 (86.7) | 13 (81.3) | 26 (89.7) |  | 1 (ref) |
| 1x | 5 (11.1) | 3 (18.8) | 2 (6.9) | 0.259 | 3.00 [0.45-20.24] |
| 2x | 1 (2.2) | 0 | 1 (3.4) | 1.000 | .. |
| **Vaccination status ≥ 12 years** |  |  |  |  |  |
| Unvaccinated | 5 (2.7) | 2 (3.8) | 3 (2.3) |  | 1 (ref) |
| 1x | 3 (1.6) | 1 (1.9) | 2 (1.5) | 0.851 | 0.75 [0.04-14.97] |
| 2x | 78 (42.4) | 15 (28.3) | 63 (48.1) | 0.282 | 0.36 [0.06-2.33] |
| 3x/booster vaccinated | 98 (53.3) | 35 (66.0) | 63 (48.1) | 0.846 | 0.83 [0.13-5.23] |
| **Antibody titer day 1, median (IQR)<sup>h</sup>** | 5000 (2000-14000) | 5000 (1000-11250) | 5500 (2000-16000) | 0.110 | 0.98 [0.96-1.00] |
| Data are presented as No. (%) unless otherwise indicated. |  |  |  |  |  |
| Abbreviations: BMI, body mass index; CI, confidence interval; IQR, interquartile range; OR, odds ratio; y, year. |  |  |  |  |  |
| <sup>a</sup> P-value for comparison between household members and index cases. |  |  |  |  |  |
| <sup>b</sup> Odds Ratio for comparison between household members and index cases. |  |  |  |  |  |
| <sup>c</sup> BMI categories for index cases and household members <18 years of age were defined as BMI z-score <-2, underweight; -2 to 1, normal weight; 1-2, overweight; >2, obesity.<sup>30</sup> BMI categories for index cases and household members ≥18 years of age were defined as <18.5 kg/m2, underweight; 18.5–24.9 kg/m2, normal weight; 25.0–29.9 kg/m2, overweight; ≥30.0 kg/m2, obesity. |  |  |  |  |  |

|  |  |
| --- | --- |
| <sup>d</sup> | Cardiovascular disease, lung disease, immune disorder, diabetes and other disorders. |
| <sup>e</sup> | Only previous infection |
| <sup>f</sup> | Only previous vaccination |
| <sup>g</sup> | Previous infection and vaccination (at least one vaccination) (hybrid immunity) |
| <sup>h</sup> | Blood available for 66 index cases (66/70, 94.3%) and 152 household members (152/171, 88.9%). |

**Table II.** Household characteristics (N=67)

| **Table II - Household characteristics (N=67)** |  |  |  |  |  |
| --- | --- | --- | --- | --- | --- |
|  | **Total households** | **Households with**<br>**secondary transmission** | **Households without**<br>**secondary transmission** | **Household secondary**<br>**attack rate (%)** |  |
| **N** | **67** | **40** | **27** | **59.7** |  |
| **Characteristic** |  |  |  | **P<sup>a</sup>** | **OR<sup>b</sup> [95%CI]** |
| **Median household size (IQR)** | 4.0 (4.0-4.0) | 4.0 (4.0-4.0) | 4.0 (4.0-4.0) | 0.798 | 1.12 [0.47-2.66] |
| **Household size, No. of persons** |  |  |  |  |  |
| 3 | 14 (20.9) | 8 (20.0) | 6 (22.2) |  | 1 (ref) |
| 4 | 45 (67.2) | 27 (67.5) | 18 (66.7) | 0.849 | 1.13 [0.33-3.79] |
| 5 | 8 (11.9) | 5 (12.5) | 3 (11.1) | 0.806 | 1.25 [0.21-7.41] |
| Data are presented as No. (%) unless otherwise indicated. |  |  |  |  |  |
| The household secondary attack rate was 59.7%. |  |  |  |  |  |
| Abbreviations: CI, confidence interval; IQR, interquartile range; OR, odds ratio. |  |  |  |  |  |
| <sup>a</sup> P-value for comparison between households with secondary transmission and households without secondary transmission. |  |  |  |  |  |
| <sup>b</sup> Odds Ratio for comparison between households with secondary transmission and households without secondary transmission. |  |  |  |  |  |

**Table III.**
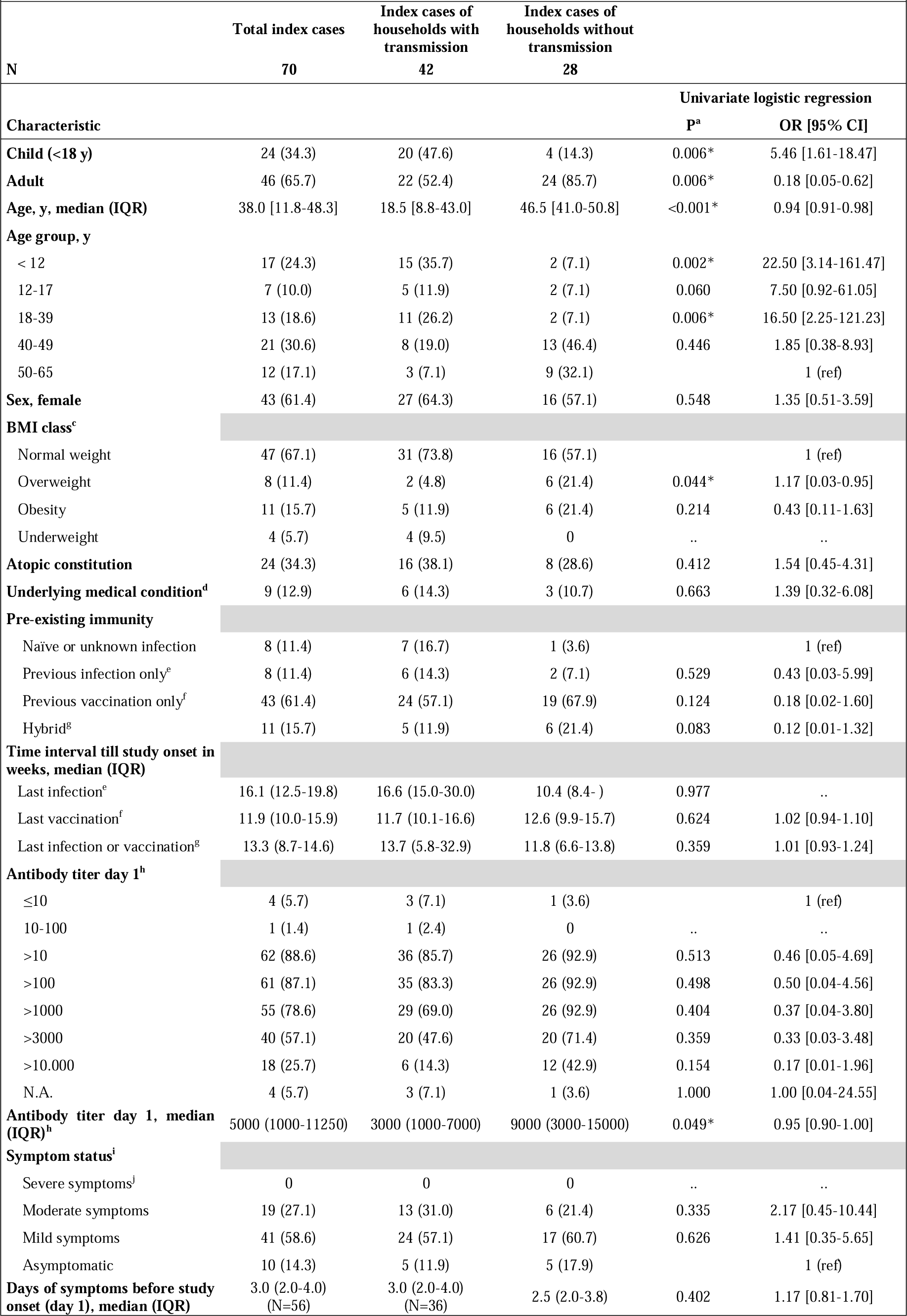
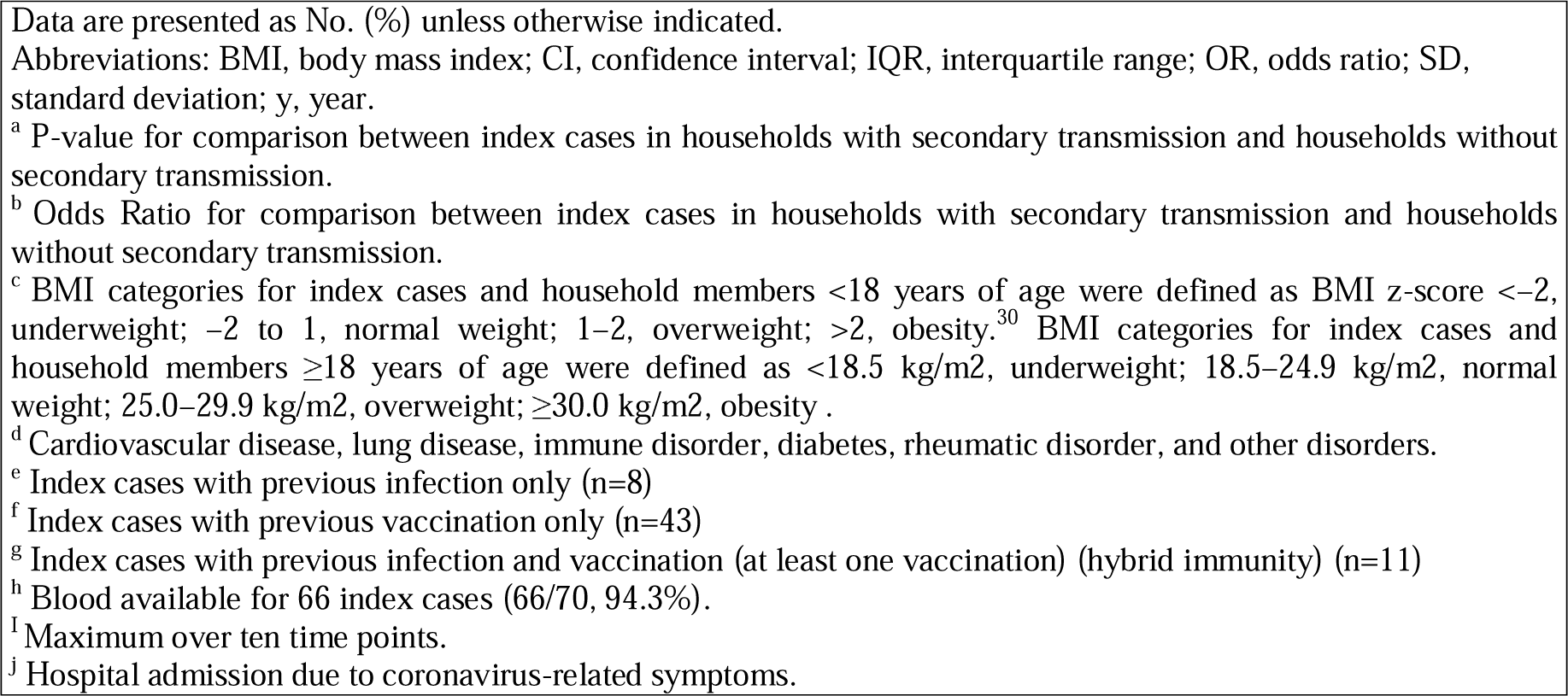
Index Case Characteristics (N=70)

| Table III – Index Case Characteristics (N=70) |  |  |  |  |  |
| --- | --- | --- | --- | --- | --- |
|  | Total index cases | Index cases of households with transmission | Index cases of households without transmission |  |  |
| N | 70 | 42 | 28 |  |  |
| Characteristic |  |  |  | Univariate logistic regression |  |
|  |  |  |  | P<sup>a</sup> | OR [95% CI] |
| Child (<18 y) | 24 (34.3) | 20 (47.6) | 4 (14.3) | 0.006* | 5.46 [1.61-18.47] |
| Adult | 46 (65.7) | 22 (52.4) | 24 (85.7) | 0.006* | 0.18 [0.05-0.62] |
| Age, y, median (IQR) | 38.0 [11.8-48.3] | 18.5 [8.8-43.0] | 46.5 [41.0-50.8] | <0.001* | 0.94 [0.91-0.98] |
| Age group, y |  |  |  |  |  |
| < 12 | 17 (24.3) | 15 (35.7) | 2 (7.1) | 0.002* | 22.50 [3.14-161.47] |
| 12-17 | 7 (10.0) | 5 (11.9) | 2 (7.1) | 0.060 | 7.50 [0.92-61.05] |
| 18-39 | 13 (18.6) | 11 (26.2) | 2 (7.1) | 0.006* | 16.50 [2.25-121.23] |
| 40-49 | 21 (30.6) | 8 (19.0) | 13 (46.4) | 0.446 | 1.85 [0.38-8.93] |
| 50-65 | 12 (17.1) | 3 (7.1) | 9 (32.1) |  | 1 (ref) |
| Sex, female | 43 (61.4) | 27 (64.3) | 16 (57.1) | 0.548 | 1.35 [0.51-3.59] |
| BMI class<sup>c</sup> |  |  |  |  |  |
| Normal weight | 47 (67.1) | 31 (73.8) | 16 (57.1) |  | 1 (ref) |
| Overweight | 8 (11.4) | 2 (4.8) | 6 (21.4) | 0.044* | 1.17 [0.03-0.95] |
| Obesity | 11 (15.7) | 5 (11.9) | 6 (21.4) | 0.214 | 0.43 [0.11-1.63] |
| Underweight | 4 (5.7) | 4 (9.5) | 0 | .. | .. |
| Atopic constitution | 24 (34.3) | 16 (38.1) | 8 (28.6) | 0.412 | 1.54 [0.45-4.31] |
| Underlying medical condition<sup>d</sup> | 9 (12.9) | 6 (14.3) | 3 (10.7) | 0.663 | 1.39 [0.32-6.08] |
| Pre-existing immunity |  |  |  |  |  |
| Naïve or unknown infection | 8 (11.4) | 7 (16.7) | 1 (3.6) |  | 1 (ref) |
| Previous infection only<sup>e</sup> | 8 (11.4) | 6 (14.3) | 2 (7.1) | 0.529 | 0.43 [0.03-5.99] |
| Previous vaccination only<sup>f</sup> | 43 (61.4) | 24 (57.1) | 19 (67.9) | 0.124 | 0.18 [0.02-1.60] |
| Hybrid<sup>g</sup> | 11 (15.7) | 5 (11.9) | 6 (21.4) | 0.083 | 0.12 [0.01-1.32] |
| Time interval till study onset in weeks, median (IQR) |  |  |  |  |  |
| Last infection<sup>e</sup> | 16.1 (12.5-19.8) | 16.6 (15.0-30.0) | 10.4 (8.4- ) | 0.977 | .. |
| Last vaccination<sup>f</sup> | 11.9 (10.0-15.9) | 11.7 (10.1-16.6) | 12.6 (9.9-15.7) | 0.624 | 1.02 [0.94-1.10] |
| Last infection or vaccination<sup>g</sup> | 13.3 (8.7-14.6) | 13.7 (5.8-32.9) | 11.8 (6.6-13.8) | 0.359 | 1.01 [0.93-1.24] |
| Antibody titer day 1<sup>h</sup> |  |  |  |  |  |
| ≤10 | 4 (5.7) | 3 (7.1) | 1 (3.6) |  | 1 (ref) |
| 10-100 | 1 (1.4) | 1 (2.4) | 0 | .. | .. |
| >10 | 62 (88.6) | 36 (85.7) | 26 (92.9) | 0.513 | 0.46 [0.05-4.69] |
| >100 | 61 (87.1) | 35 (83.3) | 26 (92.9) | 0.498 | 0.50 [0.04-4.56] |
| >1000 | 55 (78.6) | 29 (69.0) | 26 (92.9) | 0.404 | 0.37 [0.04-3.80] |
| >3000 | 40 (57.1) | 20 (47.6) | 20 (71.4) | 0.359 | 0.33 [0.03-3.48] |
| >10.000 | 18 (25.7) | 6 (14.3) | 12 (42.9) | 0.154 | 0.17 [0.01-1.96] |
| N.A. | 4 (5.7) | 3 (7.1) | 1 (3.6) | 1.000 | 1.00 [0.04-24.55] |
| Antibody titer day 1, median (IQR)<sup>h</sup> | 5000 (1000-11250) | 3000 (1000-7000) | 9000 (3000-15000) | 0.049* | 0.95 [0.90-1.00] |
| Symptom status<sup>i</sup> |  |  |  |  |  |
| Severe symptoms<sup>j</sup> | 0 | 0 | 0 | .. | .. |
| Moderate symptoms | 19 (27.1) | 13 (31.0) | 6 (21.4) | 0.335 | 2.17 [0.45-10.44] |
| Mild symptoms | 41 (58.6) | 24 (57.1) | 17 (60.7) | 0.626 | 1.41 [0.35-5.65] |
| Asymptomatic | 10 (14.3) | 5 (11.9) | 5 (17.9) |  | 1 (ref) |
| Days of symptoms before study onset (day 1), median (IQR) | 3.0 (2.0-4.0) (N=56) | 3.0 (2.0-4.0) (N=36) | 2.5 (2.0-3.8) | 0.402 | 1.17 [0.81-1.70] |

Data are presented as No. (%) unless otherwise indicated. Abbreviations: BMI, body mass index; CI, confidence interval; IQR, interquartile range; OR, odds ratio; SD, standard deviation; y, year.
<sup>a</sup> P-value for comparison between index cases in households with secondary transmission and households without secondary transmission.
<sup>b</sup> Odds Ratio for comparison between index cases in households with secondary transmission and households without secondary transmission.
<sup>c</sup> BMI categories for index cases and household members <18 years of age were defined as BMI z-score <-2, underweight; -2 to 1, normal weight; 1-2, overweight; >2, obesity.<sup>30</sup> BMI categories for index cases and household members ≥18 years of age were defined as <18.5 kg/m<sup>2</sup>, underweight; 18.5–24.9 kg/m<sup>2</sup>, normal weight; 25.0–29.9 kg/m<sup>2</sup>, overweight; ≥30.0 kg/m<sup>2</sup>, obesity .
<sup>d</sup> Cardiovascular disease, lung disease, immune disorder, diabetes, rheumatic disorder, and other disorders.
<sup>e</sup> Index cases with previous infection only (n=8)
<sup>f</sup> Index cases with previous vaccination only (n=43)
<sup>g</sup> Index cases with previous infection and vaccination (at least one vaccination) (hybrid immunity) (n=11)
<sup>h</sup> Blood available for 66 index cases (66/70, 94.3%).
<sup>i</sup> Maximum over ten time points.
<sup>j</sup> Hospital admission due to coronavirus-related symptoms.

**Table IV.**
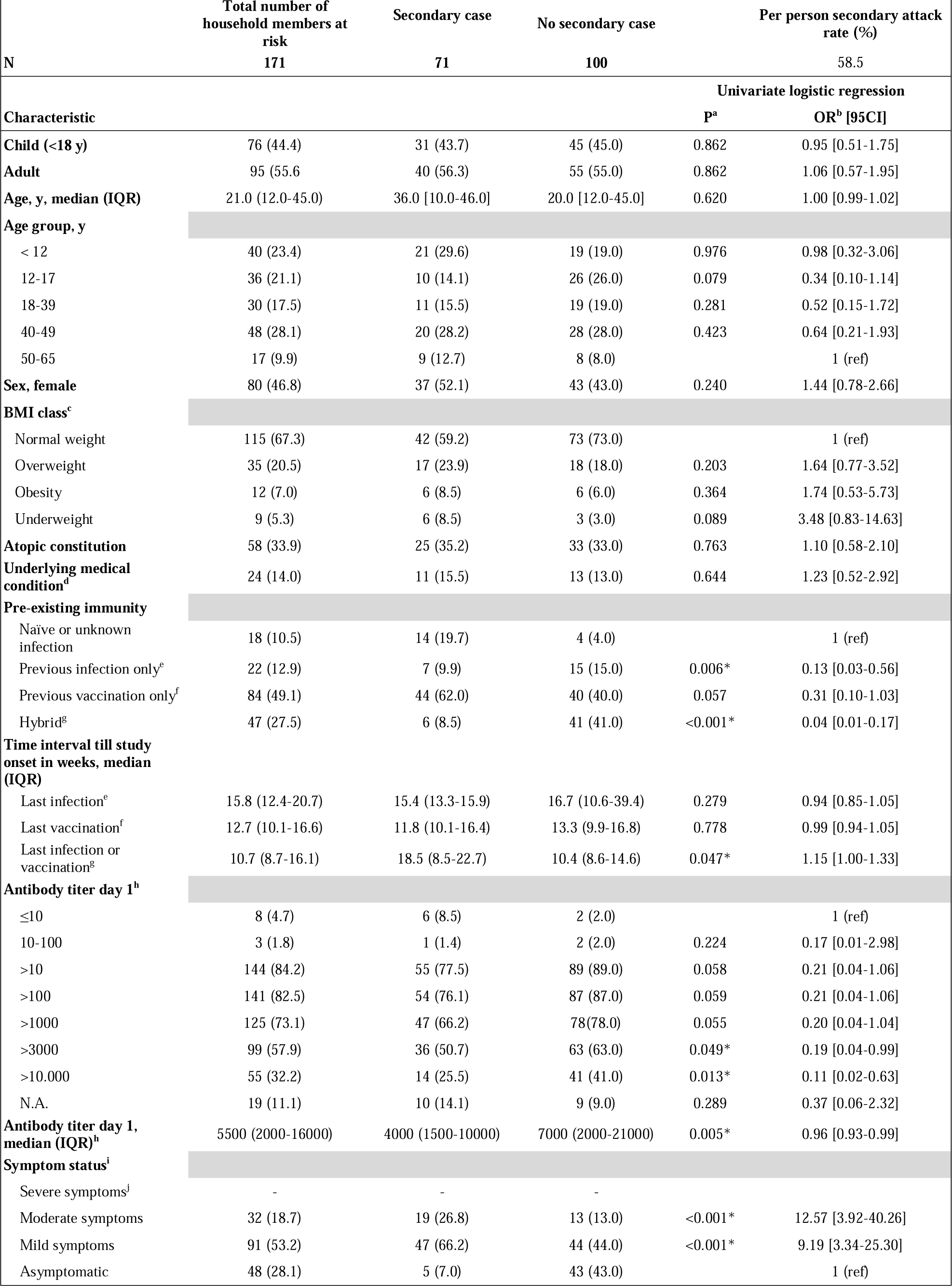
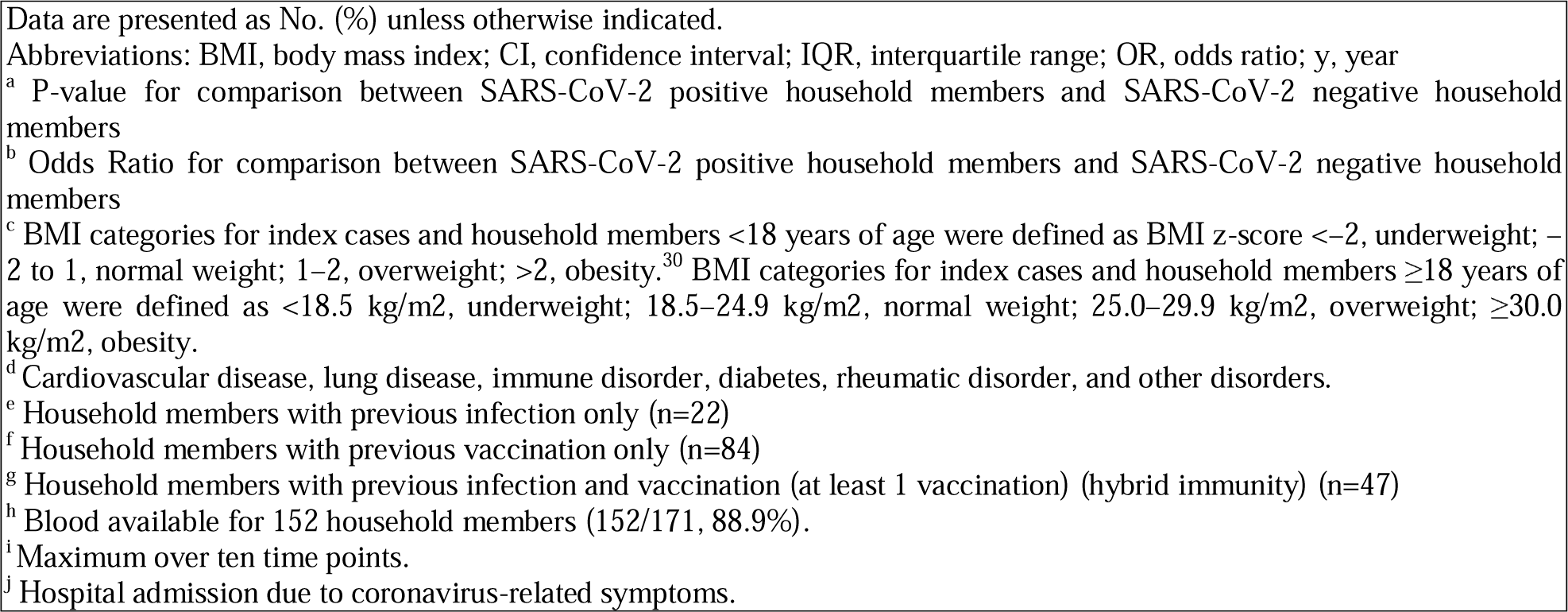
Household Member Characteristics (N=171)

Statistical analyses were performed with SPSS version 28.0.0.0. P-values <0.05 were considered statistically significant.

#### Sensitivity analysis

The household and per-person SARs calculated based on RT-PCR and serological results represent the maximum SAR, including every SARS-CoV-2 infection within a household during study follow-up. However, this method does not consider additional external introductions, leading to possible overestimation of transmission rates. A sensitivity analysis was performed to calculate a minimal household and per-person SAR, only taking into account transmission from index to household member(s) confirmed by phylogenetic analysis.

The sensitivity analysis also accounts for the influence of the index case definition on the SAR. Our defined index cases might not have been the primary cases of the household, since RPHMs could be the actual index cases of the households, therefore distorting reported household transmission dynamics.

This analysis thus corrects for 1) multiple introductions within the household as measured by whole genome sequencing and phylogenetic analysis, and 2) an RPHM being the actual index case. Then, a minimal household SAR and minimal per-person SAR were calculated. See Supplementary Methods for a detailed description of the corrections made in the sensitivity analysis.

## 3. Results

### 3.1. Baseline characteristics

Sixty-seven households were included, comprising 241 participants, of whom 70 index cases and 171 household members (Table I). In addition, 21 RPHMs were included (Supplementary Table IV). For three households, two indexes were identified. For all participants collecting specimens (n=241), protocol adherence for collection of the specimens was 93.1% (95.9% saliva and 90.2% serum) (Supplementary Table V).

Baseline characteristics of indexes and household members are shown in Table I. Most indexes were female (61.4%). Children aged <18 years accounted for 34.4% of the indexes and 44.4% of household members. Most indexes (88.6%) and household members (89.5%) had pre-existing immunity from a laboratory-confirmed SARS-CoV-2 infection, vaccination, or both.

The household characteristics are presented in Table II. The median household size was 4.0 participating household members (IQR 4.0-4.0) (Table II). The median time between index symptom onset and study enrolment (day 1) was 3.0 days (IQR 2.0–4.0) (Table III).

### 3.2. Household SAR and associations between index characteristics and SARS-CoV-2 household transmission

Secondary infection of one or more household members was detected in 40 of 67 households, leading to a household SAR of 59.7% (Table II). The median household size did not differ between households with and without transmission (4.0 [IQR 4.0–4.0] vs 4.0 [IQR 4.0–4.0]; p=0.798).

Transmission was detected in most households with index cases under the age of 18 (20/24, 83.3%) (Table III). Children had higher odds of transmitting SARS-CoV-2 to household contacts compared with adult indexes. This persisted when adjusted for baseline antibody titer (univariate OR 5.46, 95% CI 1.61–18.47, p=0.006; multivariate OR 4.75, 95% CI 1.15–19.64, p=0.031; Table III).

Other index-case characteristics, such as index symptom status or participant-reported pre-existing immunity status (infection and/or vaccination), were not statistically significantly associated with household transmission. Higher baseline antibody titer of the index case was associated with lower household transmission, however not statistically significant when correcting for age (univariate OR 0.95, 95% CI 0.90-1.00, p=0.049; multivariate OR 1.00, 95% CI 0.92-1.02, p=0.237).

### 3.3. Per-person SAR and associations between household member characteristics and SARS-CoV-2 infection

At the household member level, secondary infection was detected in 71 of 171 individuals, leading to a per- person SAR of 41.5% (Table IV). Most secondary cases already tested positive in saliva at day 1 (42/71, 61.8%). Secondary cases reported more symptoms than negative household members. The median age did not differ between secondary and non-secondary cases (36.0 [IQR 10.0–46.0] vs 20.0 [IQR 12.0–45.0]; p=0.620) (Table IV).

Household members with self-reported hybrid immunity and with previous infection only, had significantly lower odds for secondary infection compared to naïve members (OR 0.04 [95% CI 0.01–0.17]; p <0.001) vs (OR 0.13 [95% CI 0.03–0.56]; p=0.006). Previous vaccination only did not significantly lower the odds for secondary infection, independent from the number of prior vaccinations.

To analyse the association between the immune status of household members and secondary infection more specifically, members were grouped based on baseline antibody titer. Household members with high antibody titers (>3000) at day 1 had significantly lower odds for secondary infection compared to naïve members (antibody titer ≤10) (Table IV). Moreover, the median day 1 titer of non-infected household members was significantly higher than that of infected members (median 7000 vs. 4000, p=0.005). This persisted when adjusting median antibody titer for age (univariate OR 0.96, 95% CI 0.93-0.99, p=0.005; multivariate OR 0.96, 95% CI 0.93–0.99, p=0.003; Table IV).

### 3.4. Sequencing and phylogenetic results

Sequence analysis was successful for 115 SARS-CoV-2 positive individuals originating from 62 households, of whom 59 indexes and 56 household members. For 27 of 40 (67.5%) households with transmission or positive household member(s), sequences were available for both the index and all positive members. For four households, sequences were available for the index and one positive household member.

The phylogeny of these sequences showed a distinct cluster with minimal sequence differences in 23 of the 27 (85.2%) households with fully known sequences and in all four incomplete households, indicative of a single introduction within each household (Figure II). However, in four households, sequences showed large genetic distances between each of the household members or their appointed index case, indicating multiple, independent introductions of SARS-CoV-2 into the household. The subsequent sensitivity analysis took these cases into account.

### 3.5. Recent positive household members (RPHMs) and sequencing

Sequencing was performed on available isolates of RPHMs who tested positive within two weeks before inclusion to investigate whether these cases were related to the later transmission within the household. Of the six RPHMs that tested positive within two weeks before inclusion, four sequences became available. Three of these isolates showed clustering with other members of the household, including the index case, indicative of mutual transmission. One sequence was not closely related to other members, showing an independent introduction. Therefore, the three RPHMs with confirmed similar sequences could be appointed as the actual index case. Moreover, the two RPHMs with unknown sequences could not be excluded as the actual index. The subsequent sensitivity analysis corrected for these five cases.

### 3.6. Sensitivity analysis

The selection of households and household members for sensitivity analysis to calculate the minimal household and per-person SAR is visualized in Supplementary Figure I and II. First, the sensitivity analysis excluded the above-mentioned five households containing five indexes and their 13 household members with a possible other primary case (RPHM) than the defined index.

Moreover, for the calculation of the minimal household SAR, 11 additional households were excluded because transmission from the index to at least one household member could not be confirmed by sequencing (Figure II) or sequencing showed more than one introduction into the household. Transmission was confirmed by phylogenetic analysis in 26 of 51 remaining households, resulting in a minimal household SAR of 51.0%. Thereby, the household SAR estimates changed from the crude estimation (maximal household SAR) of 59.7% to the corrected sensitivity estimation (minimal household SAR) of 51.0%.

Similarly, for the calculation of the minimal per person SAR, SARS-CoV-2 positive household members were excluded if transmission from the index to this household member was not confirmed by sequencing, leading to the exclusion of 28 positive household members. Transmission from index to household member was proven by sequencing in 37 of 71 positive household members (52.1%). Since 93 negative household members were remaining, the corrected minimal per person SAR is 28.5% (37/130). Thereby, the household member SAR estimates changed from the crude estimation (maximal per-person SAR) of 41.5% to the corrected sensitivity estimation (minimal per-person SAR) of 28.5%.

The characteristics of households, index cases and household members included in the sensitivity analysis are presented in Supplementary Table I-III. Characteristics associated with transmission were not different between the primary and sensitivity analysis (Supplementary Tables I–III).

## 4. Discussion

This study prospectively analysed household transmission of the SARS-CoV-2 Omicron BA.2 VOC using dense saliva sampling and phylogenetic analysis and compared results with pre-immunisation household transmission rates of the ancestral variant, using a largely identical study design. Despite the finding that infection-acquired immunity, hybrid immunity and higher baseline antibody titer of household members significantly decreased infection risk, this study still found high transmission rates: household SAR 51.0%–59.7% and per-person SAR 28.5%-41.5%. Here, we showed a decrease of 10 percentage points for the household SAR and 14 percentage points for per-person SAR when correcting for our index case definition and infections from outside the household. The multiple simultaneous introductions detected within households emphasise the relevance of phylogenetic analysis to validate transmission within each household. This study also reveals that transmission occurs within a short time interval after infection of the index case. Moreover, underage index cases were more likely to transmit the virus to household members compared to adult indexes.

This study found high SARs, which fall within the broad range of household SARs reported in other Omicron household transmission studies, varying between 15.0%–70.2%.^8,10,22^ This wide range of reported transmission rates, partially caused by variability in study methods, has been found in all pandemic phases, comprising diverse SARS-CoV-2 variants and immune statuses.^8^ This makes comparison of SARs between pandemic phases challenging. However, comparing transmission rates is necessary to specify the net impact of the highly transmissible Omicron variant versus immunity on transmission. These challenges were overcome by comparing two studies with almost identical designs, thereby distilling the net impact. In our previous identical study performed in December 2020-January 2021, we reported household SAR of 88.2% and per-person SAR of 64.3%. The Omicron BA.2 household transmission rates observed in the current study were clearly lower compared to that observed in the initial early-pandemic study of ancestral SARS-CoV-2 in an immune naïve population:^9^ the household SAR estimates decreased from 88.2% to 59.7%, and the per-person SAR reduced from 64.3% to 41.5% compared to the initial estimates at the start of the pandemic. This remarkable decrease in household transmission rate of the Omicron VOC in a high-immunised cohort, despite the higher transmissibility phenotype of Omicron, emphasises the relevance of immunity to hamper transmission. In the current study, higher baseline antibody titer in household members was associated with protection against transmission. Thereby, participant-reported infection-induced and hybrid immunity of household members are both associated with protection against Omicron VOC household transmission, as shown in the current study and a previous household study.^10^ The vaccine effectiveness against household transmission in absence of prior infection is less prominent in this study, possibly related to the fact that vaccines contained the ancestral strain at the time or the limited size of our study population. However, various studies indicate that indexes with prior immunity acquired by vaccination reduce Omicron household transmission compared to non-vaccinated indexes,^15,17,23,24^ although vaccine effectiveness is lower for the Omicron variant than for previous variants.^25^ Thus, pre-existing immunity has a reductive impact on household transmission, although new variants challenge this by immune escaping strategies.^5^ These strategies oppose a steep reduction in transmission rate, which still allows the virus to spread substantially.

This study found that children were more likely to transmit the Omicron BA.2 variant to household members, and that children were as susceptible to infection with this variant as adult household members. However, our previous study did not find children to have a pivotal role in the household transmission of SARS-CoV-2.^9^ This shift is in line with literature, showing an increased role of children in household transmission since the emergence of VOCs compared to the first pandemic phase.^26^ One cause of increased transmission from children to their household members relative to the ancestral virus could be the different immunisation rates between children and adults, since immunisation is shown to play a role in preventing SARS-CoV-2 household transmission in this and other studies.^15,17,24,25^ However, this study observed that children still have a significantly increased risk of transmitting SARS-CoV-2 after adjusting for immunisation status. This may be because young children are more likely to spend time with family compared to other age groups. However, it also supports the hypothesis that the evolution of the virus over time resulted in an increased role for children in transmission over the course of the pandemic.^26^ This could be attributed to the finding that the ancestral strain, but not the Omicron variant, is less efficient at replicating in the primary nasal epithelial cells of children, which may impact the viral shedding of children compared to adults. ^27^

This study detected multiple introductions in at least four households by whole genome sequencing. In times of high incidence and regular social interaction patterns, it is possible that household members independently become infected by different sources. These introductions from external sources into the household may result in an overestimation of the household transmission rate if left undetected. In the initial early-pandemic study, performed in a lockdown period with strict preventative measures such as the closure of public locations and schools, sequencing and phylogenetic results showed no evidence of multiple introductions within households.^9^ To our knowledge, this is the first household transmission study performing whole genome sequencing in both indexes and household members during the Omicron wave, describing multiple introductions within one household and performing a sensitivity analysis to calculate corrected (minimal) transmission rates. Thereby, the reported transmission estimates in this study more accurately approach the true rates.

There are some limitations. First, a selection bias is introduced towards symptomatic index cases since symptomatic individuals are more likely to test themselves, while many asymptomatic cases are easily overlooked.^28^ Not only does this affect the entire pool enrolled in this study, but this might also affect the appointment of the index within households towards a symptomatic member. Although overlooking an asymptomatic index case cannot be prevented entirely, testing history and symptoms up to 8 weeks before inclusion were investigated. Second, as no lockdown measures were implemented in the Netherlands at the time of this study, and many of the participants were positive at study start, the possibility of an external case that infected all household members at once cannot be excluded. Third, recruiting after voluntary testing at facilities may introduce a volunteer bias towards those who are more inclined to adhere to COVID-19 testing measures and isolation policies.^29^ This could potentially result in higher transmission rates in the general population.

In conclusion, by using dense saliva sampling and phylogenetic analysis, this study reveals that the household transmission of the Omicron BA.2 VOC is high despite immunization. However, the transmission rate is lower compared to previous studies with other SARS-CoV-2 variants, highlighting the effect of immunity. Children have a crucial role in household transmission, while immunity has been demonstrated to provide protection against household member infection. Moreover, this study found multiple unique introductions in households by whole genome sequencing, which emphasises the importance of sequencing in future household transmission studies. These findings thereby contribute to the design of future transmission studies and the development of future public health strategies.

## Authors’ contributions

SvL, DE, MAvH, JSP, AW, MH, CR contributed to the conception and design of the study; AW, SvL, MEH, participated in the acquisition of data; DE and EK coordinated the laboratory analyses; AW, EK, SvL, JGCS, SE, DE, and MAvH were responsible for data analyses and interpretation; AW, EK, SE, SvL and DE verified the underlying data; AW, EK, SvL, DE wrote the manuscript. All authors had full access to all the data and reviewed and approved the final version of the manuscript.

## Corresponding authors

SFL van Lelyveld; D Eggink.

## Data availability statement

Deidentified data collected for the study, including individual participant data collected during the study and a data dictionary, will be shared upon reasonable request. These requests will be discussed with all project partners (Spaarne Gasthuis and RIVM). Requests should be directed to. These requests will be reviewed and approved by the investigator and project partners based on scientific merit. To gain access, data requesters will need to sign a data access agreement. Privacy-sensitive data, which is traceable to the participant, will not be shared.

## Acknowledgements, funding and meeting(s) where the information has previously been presented

There were no funding sources. The data has not been presented in international meetings.

We thank the team of the Public Health Services (GGD) Kennemerland for providing information to the (possible) participants, the laboratory team of the National Institute for Public Health and the Environment (RIVM), including Sophie van Tol, Gert-Jan Godeke, Coralie Valle, Lisa Wijsman, Bas van der Veer, Annemarie van den Brandt, Jeroen Cremer, Sharon van den Brink, Ryanne Jaarsma, Kim Freriks, Lynn Aarts, Sanne Bos, Mansoer Elahi, Jil Kocken for the laboratory analyses and the research team of the Spaarne Gasthuis Academy, particularly Jacqueline Zonneveld, Sandra Kaamer van Hoegee, Mara van Roermund, Josseline Veldhuijzen and Yara Sijm for their hard work and efforts concerning the continuation of the SARSLIVA 2.0 study.

