## Supplemental Material for "Lower levels of household transmission of SARS-CoV-2 VOC Omicron compared to Wild-type: an interplay between transmissibility and immune status"

**Supplementary Materials**

**Content**

- Supplementary Methods	Page 2-6
  - Lockdown and isolation measures
  - Sample collection
  - Case definitions
  - Molecular diagnostics and serology
  - Protein microarray
  - Whole genome sequencing and phylogeny
  - Sensitivity analysis
- Supplementary Results	Page 7, 8
  - Index cases: SARS-CoV-2 detection in different Specimens
  - Household members: SARS-CoV-2 detection in different Specimens
  - Sensitivity analysis
- References	Page 9, 10

**Other supplementary files**

- Supplementary Tables I-V	Page 11-17
- Supplementary Figures I and II	Page 18, 19

**Supplementary Methods**

Lockdown and isolation measures

During the period of this study, the Netherlands did not institute any lockdown measures. Limited isolation measures were implemented, with participants being instructed to undergo a PCR test at the Public Health Services in the event of symptoms and/or a positive self-test. Following positive test results, a minimal isolation period of 5 days was recommended, along with a requirement of at least 24 hours without symptoms before ending of isolation. Household members were not advised to isolate unless they exhibited symptoms.^1^

Sample collection

Saliva samples were self-obtained by participants at day 1, 3, 5, 7, 10, 14, 21, 28, 35, and 42 (with day of inclusion as day 1. The participants were instructed to stop drinking, eating, teeth brushing and smoking at least 30 minutes before saliva collection and to avoid clearing their throats or coughing during saliva collection. Saliva was collected using different methods depending on the age of the participants. Participants from the age of five were instructed to spit at least 2 mL of saliva into a Genefix Saliva Collection device without buffer (Isohelix), using funnels to ensure clean collection. Children below the age of five were instructed to place two Oracol (S10) sponges (Malvern Medical Developments) in the mouth for one to two minutes and rub these against the gums, before placing these in a tube. The saliva samples were stored in the participant’s home freezer. At day 42, saliva samples were transported to the laboratory on dry ice for storage at -80 degrees Celsius (°C).

At day 1 and day 42, capillary blood samples were collected by the research team during a home visit. These blood samples were transported by post to the laboratory for serological analysis. The samples were spun down and stored at -80°C upon receival at the laboratory.

Online symptomatology questionnaires were sent to the participants each night before the pre-defined time points for saliva collection. The severity was scored per symptom on a four-item scale (‘no’, ‘mild’, ‘moderate’, ‘severe’).

Saliva (self-)sampling

In our previous study, saliva self-sampling was used to study SARS-CoV-2 household transmission.^2^ Salivary (self-)sampling is highly sensitive for near real-time monitoring of SARS-CoV-2 transmission in this setting and an attractive way for monitoring SARS-CoV-2 spreading since its non-invasive nature.^2–5^

Case definitions

Previous infection was defined as a confirmed SARS-CoV-2 infection through either PCR or antigen (self-)testing before study-onset.

A household was defined as a group of persons living in the same residence with one or more shared spaces. The first member of each household with a RT-PCR confirmed SARS-CoV-2 infection on a com­bined nasopharyngeal (NP) and oropharyngeal (OP) swab (S) in the 72 hours previous to study inclusion was defined as the index case. A household was considered to have two index cases if both household members tested positive for SARS-CoV-2 at the same moment and symptoms started simultaneously (within two hours). Household members with a confirmed SARS-CoV-2 infection (by either RT-PCR or Rapid antigen self-test within eight weeks before study-onset – and therefore not eligible to fully participate – were defined as recently positive household members (RPHM). SARS-CoV-2 infection of a household member during follow-up was defined as either at least one positive SARS-CoV-2 RT-PCR result on one of the saliva samples, or a five-fold increase of serum antibodies from day 1 to day 42, regardless of the presence of symptoms.

Household transmission, defined as SARS-CoV-2 infection of at least one household member during study follow up, is expressed in the household Secondary attack rate (SAR) and the per-person SAR. The household SAR was defined as the percentage of households with secondary transmission. The per-person SAR was defined as the percentage of secondary cases out of all participating household members.

Severity of COVID-19 disease or symptoms during study follow-up was classified with a four-degree scale per time point, derived from national and international guidelines:^6,7^ (1) no coronavirus-related symptoms, (2) mild symptoms (pharyngitis, rhinitis, mild dyspnea, mild or moderate coughing, gustatory dysfunction or olfactory dysfunction), (3) moderate symptoms (moderate or severe dyspnea, severe coughing, temperature > 38°C or a pneumonia diagnosed by a physician), and (4) hospital admission due to coronavirus-related symptoms. A maximum severity score over all time points was calculated per participant.

Molecular Diagnostics and Serology

The initial NPS/OPS of index cases were tested for the presence of SARS-CoV-2 using RT-PCR by the Regional Public Health Laboratory Kennemerland, Haarlem, The Netherlands. Samples were lysed before performing RT-PCR on the SARS-CoV-2 E-gene to get a crossing point: the Cycle threshold value (Ct-value). The Ct-value was calculated using Lightcycler 480 1.5.1 software (Roche diagnostics, Basel, Switzerland).

RT-qPCR is performed on the extracted RNA of self-sampled saliva by the laboratory of the National Institute for Public Health and the Environment (RIVM), Bilthoven, the Netherlands. The extraction was executed using MagNApure 96 (MP96) with total nucleic acid kit small volume (Roche). The Roche LC480 II thermal cycler is used for PCR on 5 µl of nucleic acid in TaqMan® Fast Virus 1-Step Master Mix (Thermo Fisher) and SARS-like beta coronavirus (Sarbeco) specific E-gene primers are added and 123 probe and EAV are used as controls, as described earlier.^2^

Protein microarray

Sera were tested for the presence of immunoglobulin G antibodies reactive with the SARS-CoV-2 spike trimer and Nucleocapsid antigens in a protein microarray, in duplicate 2-fold serial dilutions starting at 1:20, essentially as described previously.^8^ For each antigen, a 4-parameter log logistic calibration curve was generated and effective concentration 50, mid-point antibody titers were calculated. Maximal fluorescent signal readout was at 43740 and minimum at 10. Raw data were processed with R version 4.04 statistical software.

Whole genome sequencing and phylogeny

Amplicon-based SARS-CoV-2 sequencing was performed on the positive saliva sample with the highest viral load for each individual using the Nanopore protocol “PCR tiling of COVID-19 virus (version: PTC_9096_v109_revE_06FEB2020),” based on the ARTIC version 3 amplicon sequencing protocol with modifications as indicated in the supplementary materials of our previous article.^2,9,10^ Purifications with AMPure XP beads and sequencing was only performed on those samples with Ctvalues below 32. Samples with Ct-values below 19 are diluted to above 19. A variable number of PCR cycles are performed before sequencing on a R9.4.1 flow cell.

The SARS-CoV-2 sequences are curated and kept based on a cutoff of 10% ambiguous nucleotides. The remaining sequences are aligned with MAFFT (v. 7).^11^ A reviewed alignment formed the base for our phylogeny built with IQtree (v. 1.6.12),^12^ using a maximum likelihood approach and TIM+F model, in accordance with the embedded ModelFinder tool.^13^ The phylogeny is rooted on the SARS-CoV-2 isolate Wuhan-Hu-1 (GenBank: NC_045512, 2020). RPHM are separately evaluated and excluded from the final phylogeny. The final tree estimation is based sequences of 115 individuals from 62 different households, but single transmissions within the household are excluded from the visualization (85 individuals from 32 families shown). The visualization is made in R (v. 4.2.2) using the ggtree library in ggplot2.^14^

A maximum likelihood phylogenetic tree is built to display clustering of sequences (Figure II). Clustering of sequences means that these sequences are within close proximity of each other in the tree, which represents a small genetic diversity between those sequences. These samples are likely very closely related or even identical, which is expected in case of actual transmission. Thereby, the proximity of sequences of household members in our tree distinguishes actual transmission within a household from an external introduction. Based on this analysis, we could correct for external introductions in our sensitivity analysis to calculate a minimal SAR.

Sensitivity analysis

The household SAR and per-person SAR calculated based on RT-PCR and serological results represent the maximum secondary attack rate, including every SARS-CoV-2 infection within a household during study follow up. However, by using this method for calculation of SAR, external introductions are not taken into account, leading to possible overestimation of transmission rates within the household. A sensitivity analysis was performed to calculate a minimal household SAR and a minimal per-person SAR only taking into account transmission which is proved by phylogenetic analysis.

The sensitivity analysis also accounts for the influence of the index case definition on the household SAR. Our defined index cases might not have been the primary case of the household, since RPHM’s could be the actual index case of the household, therefore not representing the actual household transmission dynamics.

This analysis thus corrects for 1) multiple introductions within the household as measured by whole genome sequencing and phylogenetic analysis, and 2) an RPHM being the possible actual index case.

First, we corrected the SARs for index cases that might not have been the primary case of the household, by excluding households in which it was uncertain whether the index case was the primary case. These possible primary cases are included in this study as the RPHMs. All RPHMs with a RT-PCR–confirmed SARS-CoV-2 infection 1–14 days prior to the index case could be actual index cases, based on SARS-CoV-2 incubation time and isolation rules proposed by various governments.^15^ Therefore, NP/OP-swabs of RPHMs obtained within 14 days prior to index-case SARS-CoV-2 test were sequenced. This led to the exclusion of the household of RPHMs in the sensitivity analysis if (1) the isolate of the RPHM showed clustering with the index case, implying that the RPHM would be the actual index case, or (2) the sequence of the isolate of the RPHM was unknown and the case could not be excluded as the actual index case. Households in which the sequence of the RPHM was not related to the index case were included in the sensitivity analysis, since the RPHM was assumed to be not involved in household transmission.

Second, we corrected for the possibility of multiple independent SARS-CoV-2 introductions within a household by only including households with ‘proven’ transmissions by sequencing results in our household SAR calculation. Proven household transmission is defined as the presence of at least one SARS-CoV-2 positive household member with the same, clustering viral sequence as the index case. Therefore, for the calculation of the minimal household SAR, additional households with SARS-CoV-2 positive household members were excluded if transmission from index case to at least one household member could not be proven by sequencing. This was the case if (1) index case sequence was unknown or (2) sequencing of SARS-CoV-2 positive household member(s) was unknown or had an independent acquisition from its’ index-case sequence.

**Supplementary Results**

Index cases: SARS-CoV-2 detection in different Specimens

All index cases tested positive for SARS-CoV-2 at initial NP/OP performed at Public Health Services. Of these index cases, 92.9% tested positive in at least 1 saliva sample (65/70) and 70.0% for significant increase in SARS-CoV-2 antibodies in day 42 serum specimen compared to day 1 (49/70 index cases). Three index cases did not test positive for SARS-CoV-2 in saliva nor in serum antibodies (3/70, 4.3%).

Household members: SARS-CoV-2 detection in different Specimens

Of the positive household members, 91.5% tested positive for SARS-CoV-2 in at least 1 saliva specimen (65/71) and 76.1% for a significant increase in SARS-CoV-2 antibodies in day 42 serum specimen compared to day 1 (54/71 secondary cases; Supplementary Table IV). Seventeen household members tested positive for SARS-CoV-2 in saliva only (17/71, 23.9%). Six household members tested positive for SARS-CoV-2 antibodies in serum only (6/71, 8.5%) and 48 household members tested positive for SARS-CoV-2 in saliva and serum antibodies (48/71, 67.6%).

Sensitivity analysis

First, the sensitivity analysis excluded five households containing five indexes and their 13 household members with a possible other primary case (RPHM) than our defined index. Of these 13 household members, six were SARS-CoV-2 positive and seven remained negative during study follow-up.

Moreover, for the calculation of the minimal household SAR, 11 additional households were excluded because transmission from the index case to at least one household member could not be confirmed by sequencing. This was the case if (1) the index case’s sequence was unknown or, (2) the sequence(s) of the SARS-CoV-2 positive household member(s) was unknown, or (3) the sequence(s) of the positive household member(s) was different than index case’s sequence (Supplementary Methods and Figure III). Transmission was confirmed in 26 of 51 remaining households, resulting in a minimal household SAR of 51.0%.

Similarly, for the calculation of the minimal per person SAR, SARS-CoV-2 positive household members were excluded if transmission from the index to this household member was not confirmed by sequencing, leading to the exclusion of 28 positive household members. Transmission from index case to the household member was proven by sequencing in 37 of 71 positive household members (52.1%). Since 93 negative household members remained, the corrected minimal per person SAR is 28.5% (37/130). Characteristics of index cases, households, or household members associated with secondary transmission were not different between the primary and sensitivity analysis (Supplementary Tables I–III).

16.	Centers for Disease Control Prevention (CDC). Z-score Data Files [Internet]. Available from: <https://www>.cdc.gov/growthcharts/zscore.htm

| **Supplementary Table I – Household characteristics sensitivity analysis (n=51)** | | | | | |
| --- | --- | --- | --- | --- | --- |
|  | **Total households** | **Households with secondary transmission** | **Households without secondary transmission** |  | **Secondary attack rate (%)** |
| **N** | **51** | **26** | **25** |  | 51.0% |
| **Characteristic** |  |  |  | **p^a^** | **OR^b^ [95%CI]** |
| **Median household size (IQR)** | 4.0 [3.0-4.0] | 4.0 [3.0-4.0] | 4.0 [3.5-4.0] | 0.834 | 0.90 [0.33-2.44] |
| **Household size, no of person** |  |  |  |  |  |
| 3 | 13 (25.5) | 7 (26.9) | 6 (24.0) | - | 1 (ref) |
| 4 | 34 (66.7) | 17 (65.4) | 17 (68.0) | 0.814 | 0.86 [0.24-3.09] |
| 5 | 4 (7.8) | 2 (7.7) | 2 (8.0) | 0.893 | 0.86 [0.09-8.08] |
| Data are presented as No. (%) unless otherwise indicated.  The minimal household secondary attack rate was 50.0%.  Abbreviations: CI, confidence interval; IQR, interquartile range; OR, odds ratio.  ^a^ P-value for comparison between households with secondary transmission and households without secondary transmission.  ^b^ Odds Ratio for comparison between households with secondary transmission and households without secondary transmission. | | | | | |

| **Supplementary Table II – Index case characteristics sensitivity analysis (n=52)** | | | | | |
| --- | --- | --- | --- | --- | --- |
| **No. (%)** | | | | | |
|  | **Total index cases** | **Index cases of households with secondary transmission** | **Index cases of households without secondary transmission** |  |  |
| **N** | **53** | **27** | **26** |  |  |
|  |  |  |  | **Univariate logistic regression** | |
| **Characteristic** |  |  |  | **p^a^** | **OR^b^ [95%CI]** |
| **Child (<18 y)** | 18 (34.0) | 14 (51.9) | 4 (15.4) | 0.008* | 5.92 [1.61-21.86] |
| **Adult** | 35 (66.0) | 13 (48.1) | 22 (84.6) | 0.008* | 0.17 [0.05-0.62] |
| **Age, median (IQR)** | 41.0 (11.0-48.5) | 17.0 (8.0-45.0) | 45.0 (39.5-50.3) | 0.003* | 0.95 [0.95-0.98] |
| **Age group, y** |  |  |  |  |  |
| < 12 | 13 (24.5) | 11 (40.7) | 2 (7.7) | 0.008* | 19.25 [2.18-169.79] |
| 12-17 | 5 (9.4) | 3 (11.1) | 2 (7.7) | 0.172 | 5.25 (0.49-56.80) |
| 18-39 | 8 (15.1) | 6 (22.2) | 2 (7.7) | 0.040* | 10.50 [1.12-98.91] |
| 40-49 | 18 (34.0) | 5 (18.5) | 13 (50.0) | 0.757 | 1.35 [0.21-8.82] |
| 50-65 | 9 (17.0) | 2 (7.4) | 7 (26.9) | - | 1 (ref) |
| **Sex, female** | 34 (64.2) | 19 (70.4) | 15 (57.7) | 0.338 | 1.74 [0.56-5.42] |
| **BMI class^c^** |  |  |  |  |  |
| Normal weight | 37 (69.8) | 22 (81.5) | 15 (57.7) | - | 1 (ref) |
| Overweight | 6 (11.3) | 0 | 6 (23.1) | - | - |
| Obesity | 8 (15.1) | 3 (11.1) | 5 (19.2) | 0.266 | 0.41 [0.09-1.98] |
| Underweight | 2 (3.8) | 2 (7.4) | 0 | - | - |
| **Atopic constitution** | 17 (32.1) | 10 (37.0) | 7 (26.9) | 0.432 | 1.60 [0.50-5.13] |
| **Underlying medical condition^d^** | 8 (15.1) | 5 (18.5) | 3 (11.5) | 0.481 | 1.74 [0.37-8.18] |
| **Pre-existing immunity** |  |  |  |  |  |
| Naïve or unknown infection | 7 (13.2) | 6 (22.2) | 1 (3.8) |  | 1 (ref) |
| Previous infection only^e^ | 4 (7.5) | 2 (7.4) | 2 (7.7) | 0.224 | 0.17 (0.01-2.98) |
| Previous vaccination only^f^ | 34 (64.2) | 16 (59.3) | 18 (69.2) | 0.092 | 0.15 (0.02-1.37) |
| Hybrid^g^ | 8 (15.1) | 3 (11.1) | 5 (19.2) | 0.077 | 0.10 (0.01-1.29) |
| **Time interval till study onset in weeks, median (IQR)** |  |  |  |  |  |
| Last infection^e^ | 12.5 (9.4-15.8) | 14.7 (12.6-..) | 10.4 (8.4- ) | .. | .. |
| Last vaccination^f^ | 11.5 (10.0-16.0) | 11.1 (9.8-27.7) | 12.7 (10.6-15.8) | 0.467 | 1.03 [0.95-1.11] |
| Last infection or vaccination^g^ | 13.4 (10.6-14.4) | 13.7 (11.4-..) | 13.2 (9.5-14.07) | 0.566 | 1.18 [0.67-2.10] |
| **Antibody titer day 1^h^** |  |  |  |  |  |
| ≤10 | 3 (5.7) | 2 (7.4) | 1 (3.8) |  | 1 (ref) |
| 10-100 | 1 (1.9) | 1 (3.7) | 0 | - |  |
| >10 | 47 (88.7) | 23 (85.2) | 24 (92.3) | 0.559 | 0.48 [0.04-5.65] |
| >100 | 46 (86.8) | 22 (81.5) | 24 (92.3) | 0.536 | 0.46 [0.04-4.18] |
| >1000 | 41 (77.4) | 17 (63.0) | 24 (92.3) | 0.412 | 0.35 [0.03-4.23] |
| >3000 | 29 (54.7) | 11 (40.7) | 18 (69.2) | 0.355 | 0.31 (0.03-3.78] |
| >10.000 | 14 (26.4) | 3 (11.1) | 11 (42.3) | 0.151 | 0.14 [0.01-2.07] |
| N.A. | 3 (5.7) | 2 (7.4) | 1 (3.8) | 1.000 | 1.00 [0.03-29.81] |
| **Antibody titer day 1, median (IQR)^h^** | 5000 (1000-12000) | 2000 (0-7000) | 2500 (9000-14500) | 0.140 | 0.96 [0.91-1.01] |
| **Symptom status^i^** |  |  |  |  |  |
| Severe symptoms^j^ | 0 | 0 | 0 | - | - |
| Moderate symptoms | 13 (24.5) | 8 (29.6) | 5 (19.2) | 0.605 | 1.60 [0.27-9.49] |
| Mild symptoms | 32 (60.4) | 15 (55.6) | 17 (65.4) | 0.874 | 0.88 [0.19-4.16] |
| Asymptomatic | 8 (15.1) | 4 (14.8) | 4 (15.4) | - | 1 (ref) |
| **Days of symptoms before study onset, median (IQR)** | 3.0 (2.0-4.0) | 3.0 (2.0-4.0) | 3.0 (2.0-4.0) | 0.535 | 1.13 [0.76-1.68] |
| Data are presented as No. (%) unless otherwise indicated.  Abbreviations: BMI, body mass index; CI, confidence interval; IQR, interquartile range; OR, odds ratio; SD, standard deviation; y, year.  ^a^ P-value for comparison between index cases in households with secondary transmission and households without secondary transmission.  ^b^ Odds Ratio for comparison between index cases in households with secondary transmission and households without secondary transmission.  ^c^ BMI categories for index cases and household members <18 years of age were defined as BMI z-score <–2, underweight; –2 to 1, normal weight; 1–2, overweight; >2, obesity.^16^ BMI categories for index cases and household members ≥18 years of age were defined as <18.5 kg/m2, underweight; 18.5–24.9 kg/m2, normal weight; 25.0–29.9 kg/m2, overweight; ≥30.0 kg/m2, obesity.  ^d^ Cardiovascular disease, lung disease, immune disorder, diabetes, rheumatic disorder, and other disorders.  ^e^ Only previous infection  ^f^ Only previous vaccination  ^g^ Previous infection and vaccination (at least 1 vaccination) (hybrid immunity)  ^h^ Blood available for 49 index cases (49/52, 94.2%).  ^I^ Maximum over 10 time points.  ^j^ Hospital admission due to coronavirus-related symptoms. | | | | | |

| **Supplementary Table III – Household member characteristics sensitivity analysis (n=129)** | | | | | |
| --- | --- | --- | --- | --- | --- |
| **No. (%)** | | | | | |
|  | **Total number of household members at risk** | **Secondary case** | **No secondary case** |  | **Secondary attack rate (%)** |
| **N** | **130** | **37** | **93** |  | **28.5** |
|  |  |  |  | **Univariate logistic regression** | |
| **Characteristic** |  |  |  | **p^a^** | **OR^b^ [95%CI]** |
| **Child (<18 y)** | 55 (42.3) | 13 (35.1) | 42 (45.2) | 0.298 | 0.66 [0.30-1.45] |
| **Adult** | 75 (57.7) | 24 (64.9) | 51 (54.8) | 0.298 | 1.52 [0.69-3.35] |
| **Age, y, median (IQR)** | 22.0 (12.0-45.0) | 38.0 (12.0-45.0) | 20.0 (12.0-45.0) | 0.228 | 1.01 [0.99-1.04] |
| **Age group, y** |  |  |  |  |  |
| < 12 | 28 (21.5) | 9 (24.3) | 19 (20.4) | 0.496 | 0.63 [0.17-2.37] |
| 12-17 | 27 (20.8) | 4 (10.8) | 23 (24.7) | 0.056 | 0.23 [0.05-1.04] |
| 18-39 | 22 (16.9) | 6 (16.2) | 16 (17.2) | 0.337 | 0.50 [0.12-2.06] |
| 40-49 | 39 (30.0) | 12 (32.4) | 27 (29.0) | 0.415 | 0.59 [0.17-2.09] |
| 50-65 | 14 (10.8) | 6 (16.2) | 8 (8.6) |  | 1 (ref) |
| **Sex, female** | 57 (43.8) | 18 (48.6) | 39 (41.9) | 0.487 | 1.31 [0.61-2.82] |
| **BMI class^c^** |  |  |  |  |  |
| Normal weight | 86 (66.2) | 17 (45.9) | 69 (74.2) |  | 1 (ref) |
| Overweight | 27 (20.8) | 11 (29.7) | 16 (17.2) | 0.031* | 2.79 [1.10-7.10] |
| Obesity | 11 (8.5) | 5 (13.5) | 6 (6.5) | 0.066 | 3.38 [0.92-12.41] |
| Underweight | 6 (4.6) | 4 (10.8) | 2 (2.2) | 0.021* | 8.12 [1.37-48.06] |
| **Atopic constitution** | 43 (33.1) | 14 (37.8) | 29 (31.2) | 0.467 | 1.34 [0.61-2.98] |
| **Underlying medical condition^d^** | 19 (14.6) | 8 (21.6) | 11 (11.8) | 0.159 | 2.06 [0.75-5.61] |
| **Pre-existing immunity** |  |  |  |  |  |
| Naïve or unknown infection | 11 (8.5) | 7 (18.9) | 4 (4.3) |  | 1 (ref) |
| Previous infection only^e^ | 18 (13.8) | 3 (8.1) | 15 (16.1) | 0.015* | 0.11 [0.02-0.66] |
| Previous vaccination only^f^ | 61 (46.9) | 24 (64.9) | 37 (39.8) | 0.144 | 0.37 [1.00-1.40] |
| Hybrid^g^ | 40 (30.8) | 3 (8.1) | 37 (39.8) | <0.001* | 0.05 [0.01-0.25] |
| **Time interval till study onset in weeks, median (IQR)** |  |  |  |  |  |
| Last infection^e^ | 15.8 (11.6-26.6) | 13.3 (12.6- -) | 16.7 (10.6-39.4) | 0.442 | 0.94 (0.79-1.11) |
| Last vaccination^f^ | 12.3 (10.1-16.6) | 11.4 (10.2-15.5) | 13.3 (9.9-20.6) | 0.399 | 1.00 [0.99-1.01] |
| Last infection or vaccination^g^ | 10.8 (8.8-16.7) | 20.1 (16.9- -) | 10.3 (8.6-15.7) | 0.028* | 1.31 [1.03-1.67] |
| **Antibody titer day 1^h^** |  |  |  |  |  |
| ≤10 | 5 (3.8) | 3 (8.1) | 2 (2.2) |  | 1 (ref) |
| 10-100 | 3 (2.3) | 1 (2.7) | 2 (2.2) | 0.472 | 0.33 [0.02-6.65] |
| >10 | 113 (86.9) | 31 (83.8) | 82 (88.2) | 0.141 | 0.25 [0.04-1.58] |
| >100 | 110 (84.6) | 30 (81.1) | 80 (86.0) | 0.139 | 0.25 [0.04-1.57] |
| >1000 | 96 (73.8) | 25 (67.6) | 71 (76.3) | 0.124 | 0.24 [0.04-1.49] |
| >3000 | 77 (59.2) | 21 (56.8) | 56 (60.2) | 0.144 | 0.25 [0.04-1.60] |
| >10.000 | 44 (33.8) | 8 (21.6) | 36 (38.7) | 0.054 | 0.15 [0.02-1.04] |
| N.A. | 12 (9.2) | 3 (8.1) | 9 (9.7) | 0.183 | 0.22 [0.02-2.04] |
| **Antibody titer day 1, median (IQR)^h^** | 5000 (2000-16250) | 4000 (0-10500) | 7000 (2000-19000) | 0.034* | 0.96 [0.93-1.00] |
| **Symptom status^i^** |  |  |  |  |  |
| Severe symptoms^j^ | 0 | 0 | 0 | - | - |
| Moderate symptoms | 19 (14.6) | 8 (21.6) | 11 (11.8) | 0.003* | 28.36 [3.19-251.89] |
| Mild symptoms | 71 (54.6) | 28 (75.7) | 43 (46.2) | 0.002* | 25.40 [3.30-195.54] |
| Asymptomatic | 40 (30.8) | 1 (2.7) | 39 (41.9) |  | 1 (ref) |
| Data are presented as No. (%) unless otherwise indicated.  Abbreviations: BMI, body mass index; CI, confidence interval; IQR, interquartile range; OR, odds ratio; y, year.  ^a^ P-value for comparison between SARS-CoV-2 positive household members and SARS-CoV-2 negative household members  ^b^ Odds Ratio for comparison between SARS-CoV-2 positive household members and SARS-CoV-2 negative household members  ^c^ BMI categories for index cases and household members <18 years of age were defined as BMI z-score < –2, underweight; –2 to 1, normal weight; 1–2, overweight; >2, obesity.^16^ BMI categories for index cases and household members ≥18 years of age were defined as <18.5 kg/m2, underweight; 18.5–24.9 kg/m2, normal weight; 25.0–29.9 kg/m2, overweight; ≥30.0 kg/m2, obesity.  ^d^ Cardiovascular disease, lung disease, immune disorder, diabetes, rheumatic disorder, and other disorders.  ^e^ Only previous infection  ^f^ Only previous vaccination  ^g^ Previous infection and vaccination (at least 1 vaccination) (hybrid immunity)  ^h^ Blood available for 117 household members (117/129, 90.7%).  ^I^ Maximum over 10 time points.  ^j^ Hospital admission due to coronavirus-related symptoms. | | | | | |

| **Supplementary Table IV:** **Baseline characteristics of recent* SARS-CoV-2 positive household members (N=21)** | |
| --- | --- |
| **Characteristic** | **Recent* SARS-CoV-2 positive household member** |
| **Child (<18 years)** | 11 (52.4) |
| **Age, years**, median (IQR) | 17.0 (9.5-47.5) |
| **Sex, female** | 9 (42.9) |
| **Time between SARS-CoV-2 positive test and positive test index-case**, days, median (IQR) | 31.0 (6.0-38.0) |
| **Time between SARS-CoV-2 positive test and inclusion,** days, median (IQR) | 33.0 (8.0-40.0) |
| **Symptom status^a^** |  |
| Severe symptoms^b^ | 0 |
| Moderate symptoms | 8 (38.1) |
| Mild symptoms | 12 (57.1) |
| Asymptomatic | 1 (4.8) |
| **Symptom duration, days**, median (IQR) | 7.5 (4.3-13.5) |
| **Time between symptom stop date and SARS-CoV-2 test index-case, days**, median (IQR) | 21.0 (-0.5-29.0) |
| Data are presented as No. (%) unless otherwise indicated.  Abbreviation: IQR, interquartile range.  *Within 8 weeks prior to index GGD-test.  ^a^ Maximum over 10 time points.  ^b^ Hospital admission due to coronavirus-related symptoms. | |

| Supplementary Table V: Compliance to sampling scheme | |
| --- | --- |
| Saliva, average | **95.9%** |
| Saliva day 1 | 238/241, 98.8% |
| Saliva day 3 | 236/241, 97.9% |
| Saliva day 5 | 229/241, 95.0% |
| Saliva day 7 | 232/241, 96.3% |
| Saliva day 10 | 230/241, 95.4% |
| Saliva day 14 | 230/241, 95.4% |
| Saliva day 21 | 223/241, 92.5% |
| Saliva day 28 | 226/241, 93.8% |
| Saliva day 35 | 234/241, 97.1% |
| Saliva day 42 | 234/241, 97.1% |
| Serum, average | **90.2%** |
| Serum day 1 | 218/241, 90.5% |
| Serum day 42 | 217/241, 90.0% |
| Compliance to sampling scheme for all participants collecting specimens (n=241). Protocol adherence for collection of the specimens was 93.1% (95.9% saliva and 90.2% serum). | |

Supplementary Figure I – Selection of households for sensitivity analysis

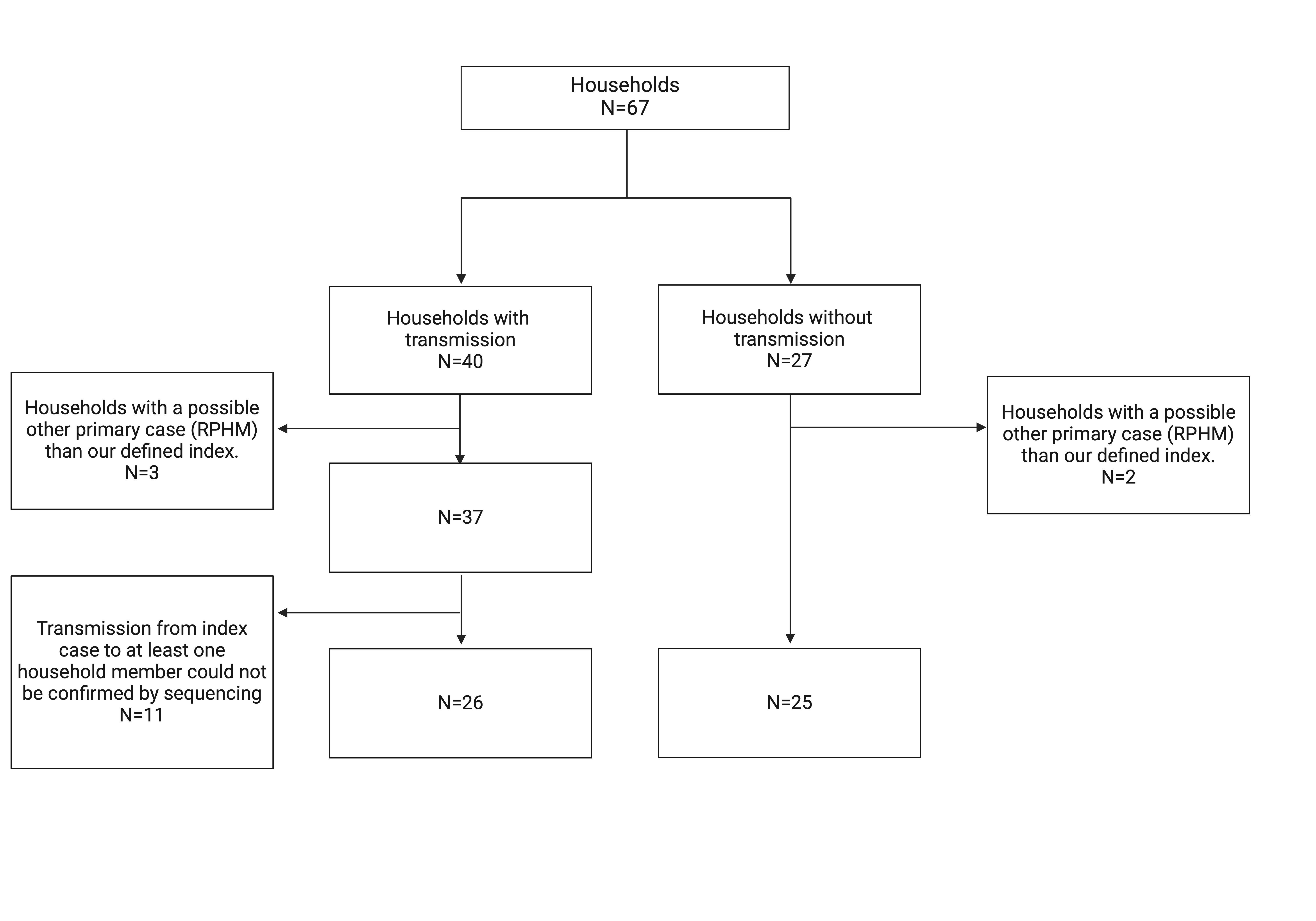

To account for the influence of our index case definition on the household SAR, and for multiple independent SARS-CoV-2 introductions within a household, a sensitivity analysis was performed based on whole genome sequencing and phylogenetic analysis. We calculated a minimal household SAR of 51.0% (26/51).

Abbreviation: RPHM, Recent positive household member

Supplementary Figure II – Selection of household members for sensitivity analysis

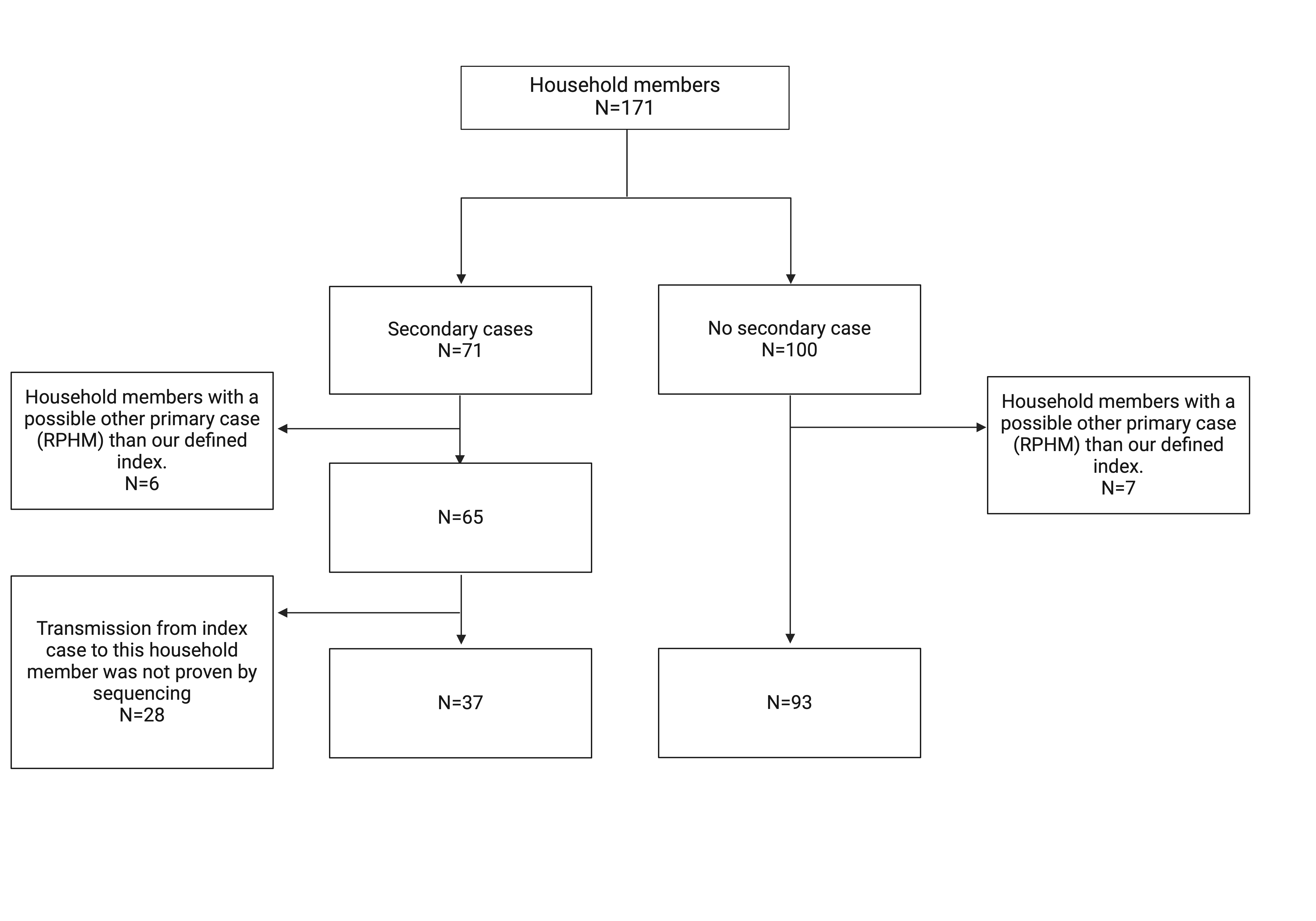

To account for the influence of our index case definition on the per-person SAR, and for multiple independent SARS-CoV-2 introductions within a household, a sensitivity analysis was performed based on whole genome sequencing and phylogenetic analysis. We calculated a minimal per-person SAR of 28.5% (37/130).

Abbreviation: RPHM, Recent positive household member
